# Changes in Global Quality of Life after treatment with immune checkpoint inhibitors in patients with advanced stage lung cancer in the Netherlands – a 2015-2021 cohort study

**DOI:** 10.1101/2024.04.26.24306439

**Authors:** Ananya Malhotra, Erick S. Zepeda, Petra C. Vinke, Geertruida H. de Bock, Willemijn Maas, T. Jeroen. N. Hiltermann, Bernard Rachet, Clémence Leyrat, Manuela Quaresma

**Affiliations:** Inequalities in Cancer Outcomes Network, Department of Health Services Research and Policy, Faculty of Public Health and Policy, London School of Hygiene & Tropical Medicine, United Kingdom; Department of Epidemiology, University Medical Center Groningen, University of Groningen, Groningen, the Netherlands; Department of Pulmonology, University of Groningen, University Medical Center Groningen, Groningen, the Netherlands; Inequalities in Cancer Outcomes Network, Department of Medical Statistics, Faculty of Epidemiology and Population Health, London School of Hygiene & Tropical Medicine, United Kingdom

**Keywords:** Quality of life, Immune checkpoint inhibitors, Joint modelling, Cancer treatments, Lung cancer

## Abstract

**Background:** Introduction of immune checkpoint inhibitors (ICIs) has modified treatment modalities for patients with lung cancer, offering new alternatives for treatment. Despite improved survival benefits, ICIs may cause side-effects impacting patients’ quality of life (QoL). We aim to study the changes in global QoL (gQoL) up to 18 months of patients with advanced stage lung cancer after treatment with ICIs between 2015-2021.

**Methods and Analysis:** A longitudinal cohort study was conducted using OncoLifeS data-biobank from the University Medical Center Groningen. Participants completed the EORTC QLQ-C30 questionnaire, at the beginning of their ICI treatment (baseline) and then at 6, 12 and 18 months. Using joint modelling, the predicted trajectory of gQoL was studied by treatment regimens up to 18 months, while accounting for the competing risk of death and adjusting for pre-specified covariates.

**Results:** Of 418 participants with median age 66 years, 39% were women. Patients receiving first line immuno-monotherapy with palliative intent had a small improvement ([5-8] points) in their gQoL within first six months, and no clinically significant change ([−5 to 5] points) thereafter, while patients with second/further line immunotherapy with palliative intent had no clinically significant change in their gQoL over 18 months. Patients receiving first line chemo-immunotherapy with palliative intent had a small improvement in their gQoL over 18 months, while patients receiving first line chemo-radiotherapy followed by Durvalumab with curative intent had no clinically significant change in their gQoL over 18 months.

**Conclusion:** The differences in gQoL over time among patients with varying treatment regimens based on drug intensity, line and intent of treatment may help clinicians and patients understand the potential dynamic of treatments on QoL. It may further influence treatment decisions and patient management strategies, reflecting the practical implications of different treatment regimens.

**Key Messages:** **What is already known on this topic**

Immunotherapy has significantly helped in modifying treatment modalities for patients with advanced stage (stage 3 and 4) lung cancer, despite the unsatisfactory prognosis of this disease. In comparison to conventional therapies like chemotherapy, various trials reported more favourable outcomes in the health related QoL domain for patients who underwent treatment with immune checkpoint inhibitors (ICIs), which is a type of immunotherapy. Despite its benefits in terms of survival, longer time until deterioration in QoL and better control of symptoms after immunotherapy, ICIs may impact patients’ QoL due to its side-effects. However, such evidence has largely been drawn from clinical trials which not only have a strict eligibility criteria but also a relatively short follow-up of under one year, while treatment with immunotherapy may last up to two years. Although some patients with lung cancer achieve deep and durable responses with ICIs, not all patients benefit and develop side-effects which may impact their QoL.

**What this study adds**

The main aim of this analyses was to study the changes in gQoL (measure by the European Organization for Research and Treatment of Cancer Quality of Life, EORTC QLQ-C30 questionnaire, version 3) over a period of 1.5 years after receiving treatment with ICIs for advanced stage lung cancer. Using joint models which account for the competing risk of death and adjusts for prespecified covariates such as baseline sociodemographic and clinical characteristics of the patients, we predicted and assessed the differences in the gQoL over time of patients who had different treatment regimens for their lung cancer diagnosis. Our analyses showed that patients receiving first line immuno-monotherapy with a palliative intent had a small improvement in their gQoL within first six months of ICI treatment, a trivial change up to one year and then a small deterioration thereafter, compared to patients with second/further line immunotherapy, who had a trivial change in their gQoL over 1.5 years after ICI treatment. Moreover, patients who had first line chemo-radiotherapy followed by Durvalumab with a curative intent had a small deterioration in their gQoL in first six months following ICI treatment, a trivial change up to one year and then a small improvement thereafter. Patients receiving first line chemo-immunotherapy with a palliative intent had a small improvement in their gQoL over 1.5 years. As a secondary outcome, we studied how the functional scores of patients change over 1.5 years, indicating their physical, social, emotional, role and cognitive wellbeing. While emotional, social and cognitive scores had trivial changes over time, physical and role functioning had a small deterioration over 1.5 years after ICI treatment.

**How this study might affect research, practice or policy**

The differences in gQoL over time among patients with varying treatment regimens based on drug combination, line and intent of treatment may help guide clinicians and patients of potential benefits and impairments of treatments on QoL.

## Introduction

Lung cancer is the leading cause of cancer-related death worldwide (about 1.8 million (18%) deaths globally in 2020^1^), largely because of high proportion of advanced-stage tumours with poor prognosis. The pattern is similar in the NL, where advanced disease represents 49% of diagnosed cases, with 1-year overall survival at 46% for patients diagnosed between 2012-2018^2^. The prognosis of advanced-stage disease is poor, with 1-year overall survival of 46% in the Netherlands for patients diagnosed between 2012-2018^2^. Despite relatively recent advances in diagnosis and therapy, the prognosis for patients with advanced-stage lung cancer is still unsatisfactory. Since 2014, introduction of immune checkpoint inhibitors (ICIs) has modified treatment modalities for patients with lung cancer, offering new alternatives for this disease in advanced stages^3^. Despite its undeniable benefit in terms of survival^4^, ICIs can cause immune-related adverse events (IRAEs), which in turn have an impact on the patients’ quality of life (QoL)^5^.

In comparison to conventional therapies like chemotherapy, various studies report smaller impairments in health related QoL scales, longer time until deterioration in QoL and better control of symptoms after immunotherapy^6–9^. This may be related to lower risk of adverse events for immunotherapy compared to chemotherapy^10^. However, this evidence has largely come from clinical trials which have strict eligibility criteria, e.g., these data exclude patients with poor performance status (Eastern Cooperative Oncology Group (ECOG) performance status (PS) >1), concomitant cancers, or comorbidities. Hence, generalising clinical benefits of ICIs seen in trial settings to real-world cohorts is hazardous.

Population-based studies have revealed that patients’ socio-demographic characteristics such as age, sex, and education, as well as health status captured via performance status, comorbidities or tumour stage may impact their QoL^11–14^. Hence, the effect of cancer treatments on QoL outcomes may be confounded by such factors. Therefore, identifying determinants of QoL is key to optimise patients’ QoL^15^.

Very few methodical studies have been published on post-treatment longer-term QoL from observational real-world data focusing on immunotherapy as well as on other cancer treatments such as chemotherapy and radiotherapy^11–13^ ^16^. Moreover, studies that investigated QoL in this population so far had a follow-up of under one year^17^, while ICI treatment regimens typically have an intended duration of two years^18^. Patients’ QoL may therefore be affected, even long after treatment with ICIs is initiated. The goals of therapy for advanced stage lung cancer should not only focus on controlling the disease but should also be directed towards optimising patient’s longer-term QoL.

This study aims to determine how global quality of life (gQoL) of patients with advanced stage lung cancer changes over 18 months following treatment with ICIs. With this knowledge, clinicians and patients can be better informed as to what to expect from a treatment with ICIs, both during and after receiving the treatment.

## Methods

### Data source

This study was based on a subset of the OncoLifeS (Oncological Life Study: Living well as a cancer survivor) data^19^, which is a hospital-based biobank of clinical well-being and QoL of patients with an oncological diagnosis and treated with anti-PD-1/PD-L1/CTLA-4 ICIs at the University Medical Center Groningen (UMCG), the Netherlands. These data consist of linked routine clinical data, including cancer treatments, comorbidities, lifestyle, radiological and pathological findings, IRAEs with patient-reported data, including QoL, that are collected during and after their cancer treatment. This biobank was developed for oncological research with an overall aim to link routine clinical data with preserved biological specimens and QoL assessments. It includes, amongst others, patients diagnosed with lung cancer and treated with ICIs from January 2015 onwards, who filled questionnaires measuring their QoL around the time of ICI initiation and then at every 6 months, up to two years. The data was pseudonymized by the project coordinator of OncoLifeS before analysis^20^.

### Inclusion criteria

The OncoLifeS database includes patients who were ≥18 years of age at the time of signing an informed consent. Patients receiving ICI treatment are those who have received one of the following monoclonal antibodies: Nivolumab, Pembrolizumab, Cemiplimab, Atezolizumab, Avelumab or Durvalumab^21^. We included patients who were diagnosed with advanced stage (3 or 4) lung cancer and treated with ICIs between January 2015 to November 2021, with no missing information on cancer treatments and who filled at least one QoL questionnaire at baseline - either at the time of ICI initiation or up to six weeks before.

### Patient and public involvement

Patients and the public were not involved in the design, conduct, reporting, or dissemination plans of this research.

### Outcomes, exposure and covariables

Our primary outcome of interest was the gQoL score of patients diagnosed with lung cancer and treated with ICIs, measured by the EORTC QLQ-C30. The European Organization for Research and Treatment of Cancer Quality of Life^22^ (EORTC QLQ-C30) questionnaire (version 3) is a part of an integrated system which provides a QoL instrument to facilitate international clinical trials in oncology. This is a 30-item questionnaire which assesses the gQoL, five functional scales (emotional, physical, social, role and cognitive), eight symptom scales (pain, fatigue, nausea/vomiting, dyspnoea, insomnia, loss of appetite, constipation, and diarrhoea) and perceived financial impact of the disease. These scales produce a continuous measure ranging from 0 to 100 with higher scores representing higher gQoL/ higher level of functioning/ higher level of symptoms. The secondary outcomes of interest were the five functional scales of QoL.

Patients were classified into four groups according to their type and line of treatment (Table 1). Hence, cancer treatment group (the exposure) was a categorical variable. These groups reflect the intensity of the treatment regimens, therefore their possible implications on the patient’s QoL, and the therapeutic intention (and indirectly patient’s prognosis).

**Table 1:**
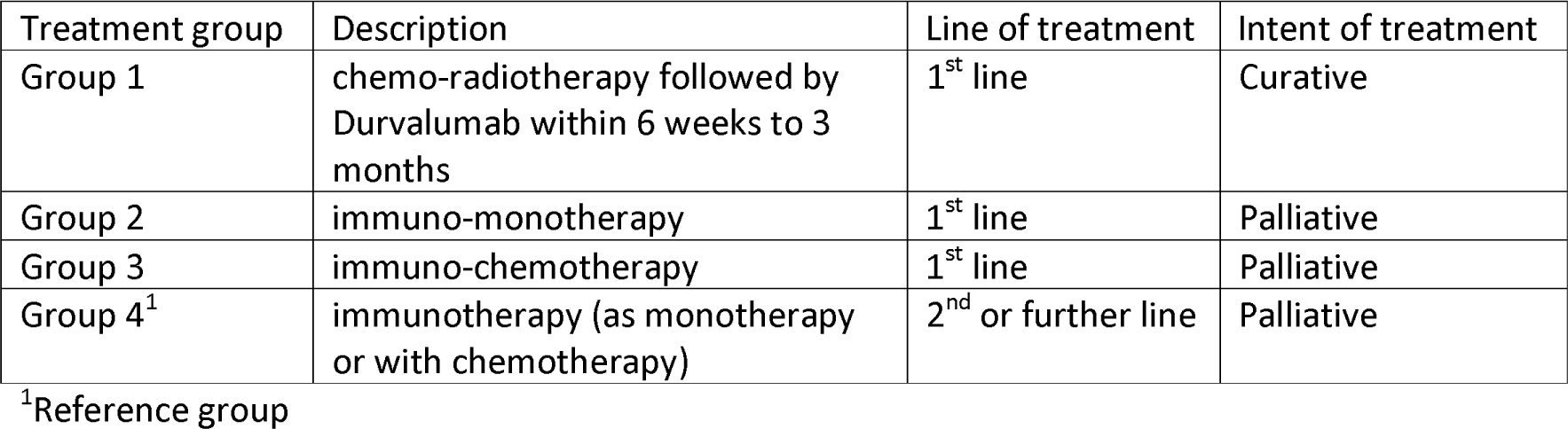
Criteria for classification of patients by their treatment regimen.

| Treatment group | Description | Line of treatment | Intent of treatment |
| --- | --- | --- | --- |
| Group 1 | chemo-radiotherapy followed by Durvalumab within 6 weeks to 3 months | 1<sup>st</sup> line | Curative |
| Group 2 | immuno-monotherapy | 1<sup>st</sup> line | Palliative |
| Group 3 | immuno-chemotherapy | 1<sup>st</sup> line | Palliative |
| Group 4<sup>1</sup> | immunotherapy (as monotherapy or with chemotherapy) | 2<sup>nd</sup> or further line | Palliative |

<sup>1</sup>Reference group

### Statistical analyses

The aim of this analysis was to study the changes in QoL of advanced stage lung cancer patients, following their ICI treatment to up to 18 months. The repeated measurements on QoL scores enabled us to study how QoL changed over time. Violin plots were used to describe the trajectories of continuous scores (i.e., gQoL and the functional scales) over time from baseline to up to 18 months, conditional on patients’ survival at each follow-up time. Violin plots could not be used for symptom scores because of the discrete nature of these sub-scores.

For assessing the longitudinal trajectory of QoL over time after ICI treatment, the competing risk of death was accounted for as it precludes patients from the outcome of interest, i.e., QoL. Joint models^23^ were used to estimate the effect of treatment on QoL over time following ICI treatment. Its two components allowed the simultaneous analysis of longitudinal and time-to-event data, which were linked using an association structure that quantifies the relationship between the changes in QoL and survival. Due to high number of patients drop-out from either death or incomplete follow-up, we restricted our analysis to up to 18 months. Models were adjusted at the time of analysis for pre-specified covariates, identified using a directed acyclic graph (DAG) created using DAGitty v3.0^24^ (Figure A.1). Baseline covariables – age, sex, weight, education, ECOG PS, presence of concomitant cancer, presence of comorbidities (diabetes, hypertension, COPD, CVD, rheumatological conditions), number of months since lung cancer diagnosis, and tumour stage were identified as adjustment factors. ICI treatment response, IRAEs and ‘ICI stopping early’ were mediators in the path between treatment and QoL and hence not adjusted for in the analysis.

The first component of the joint model was fitted using a linear mixed effect model with a patient-specific random intercept to account for the repeated measurements of QoL. The estimand was the adjusted mean difference in QoL between cancer treatments (Table1), from baseline and then every six months and up to 18 months. This was assessed by the difference between the predicted mean gQoL in the treatment groups compared above, pooled at each time point. The other component of the joint model was the time-to-event model, in the form of a proportional hazards model adjusted for the same pre-specified covariates. As a primary analysis, a complete-case analysis was performed using *JM* R package^25^ so the observations with missing data in the covariates were excluded from this analysis (Appendix 4).

Since a complete-case analysis may lead to biased results if data are not missing completely at random (MCAR) or if the missingness mechanism is not covariate-dependent only, we applied multiple imputation for handling missing data in the covariates - weight, education, and PS under a missing at random (MAR) mechanism allowing for a possible association between missingness, treatment groups, covariates, and outcome. Complex models such as joint models for longitudinal and survival data, in the presence of missing values, cannot be handled adequately by standard multiple imputation techniques. Using *JointAI* R package^26^, we fitted joint models using a fully Bayesian approach by modelling the analysis model (the joint model described above) jointly with the incomplete covariates under MAR assumption^27^, such that the analysis and imputation of missing data were performed simultaneously while ensuring compatibility between longitudinal and survival sub-models. Posterior mean difference (PM) and its 95% CI was used to describe the measure of association between gQoL and covariates. Model summaries are presented in Table A.3. To assess the differences between the trajectory of predicted mean gQoL by treatment group, we included an additional interaction term between observation time of QoL and treatment group (Figure A.13). Similarly, an additional interaction term between observation time of QoL and comorbidities was included in the model where we assessed the differences between the trajectory of predicted mean gQoL by presence of comorbidities at baseline (Figures A.14-A.16).

### Sensitivity analyses

We performed a sensitivity analysis by imputing missing observations on education under two extreme missing not at random (MNAR) mechanisms (i.e., missing information related to non-measured data), assuming in turn that all the patients with missing education had a low level of education, and then assuming they had a high level of education. We then compared the predicted trajectory of gQoL with the results obtained using multiple imputation above. We did not conduct such a sensitivity for weight and PS, as the proportion of missing data on these variables was very low.

All analyses were performed using R software version 4.3.0^28^.

## Results

There were 508 patients diagnosed with lung cancer between 1987-2021 and treated with ICIs between 2015-2021 at UMCG. Among these, 418 (82%) patients were included in our analysis because they filled at least one QoL questionnaire and were diagnosed with stage 3 or 4 lung cancer. A consort diagram was used to describe the timeline of filling EORTC QLQ-C30 questionnaires (Figure A.2). Patients’ characteristics at baseline are presented in Table 2 by treatment groups. The median age of patients at baseline was 66 years (Q1=59, Q3=71) and 161 (39%) were women. Majority patients (n=262, 63%) had second/further line immunotherapy (group 4). Most patients had stage 4 lung cancer (n=369, 88%), about half of the patients were restricted in physically strenuous activity (PS=1, n=204, 49%), while 38% had a history of CVD (n=158), 58% had low level of education (n=243). The observed median gQoL of all patients at baseline was 58.3 (Q1=50, Q3=75) (Figure 1A). The observed evolution of the gQoL of patients from baseline to up to 18 months, among those who were alive for at least 6, 12, and 18 months are shown using violin plots in Figure 1B-D respectively. The number of patients alive at follow-up times rapidly decreased, with only 53% patients alive at 12 months and 43% alive for at least 18 months. Patients who survived longer (at least 6 months) after immunotherapy had a higher gQoL at baseline and over time compared to those who died earlier (Figure 1B-D). A decline in the gQoL, physical and role functioning scores among those who survived after one year of ICI treatment was also observed (Figures 1 and A.3-A.7).

**Figure 1:**
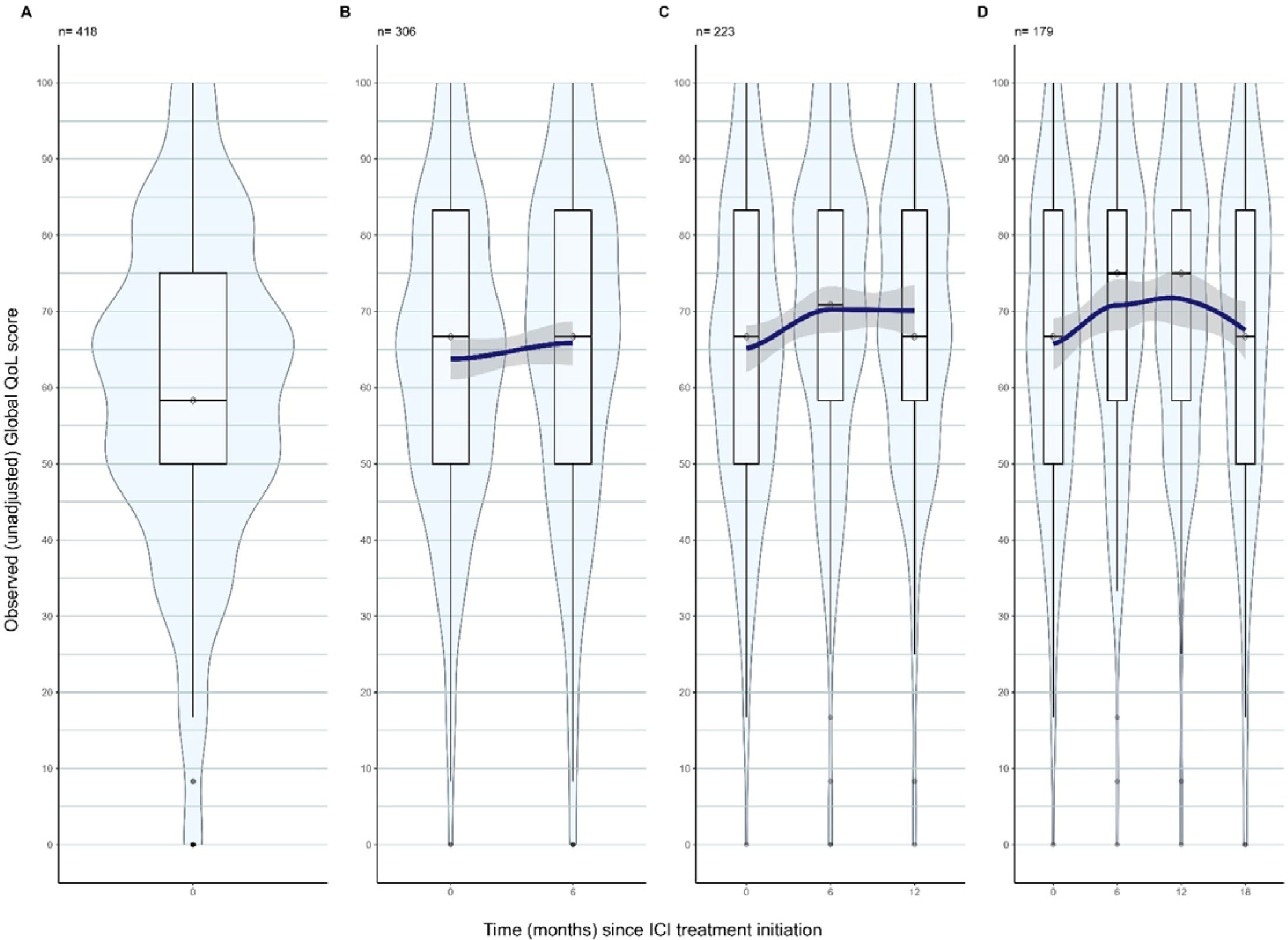
Violin plots with superimposed smoothing curve (fitted using weighted least squares) describing the observed (unadjusted) evolution of gQoL scores among patients who survived after ICI treatment, where ‘n’ is the number of patients alive at baseline (panel A), up to 6 months (panel B), up to 12 months (panel C), and up to 18 months (panel D).

**Table 2:**
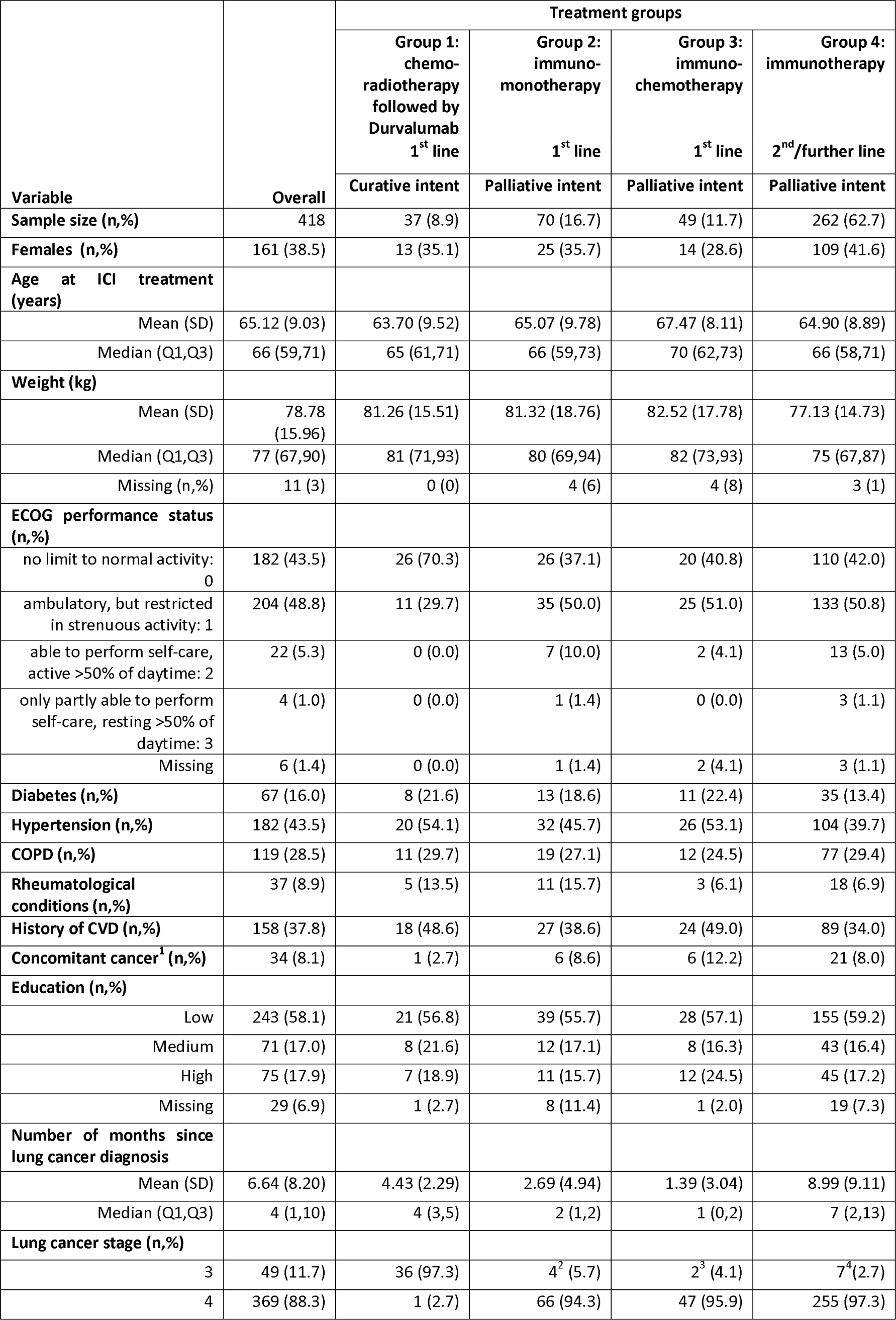
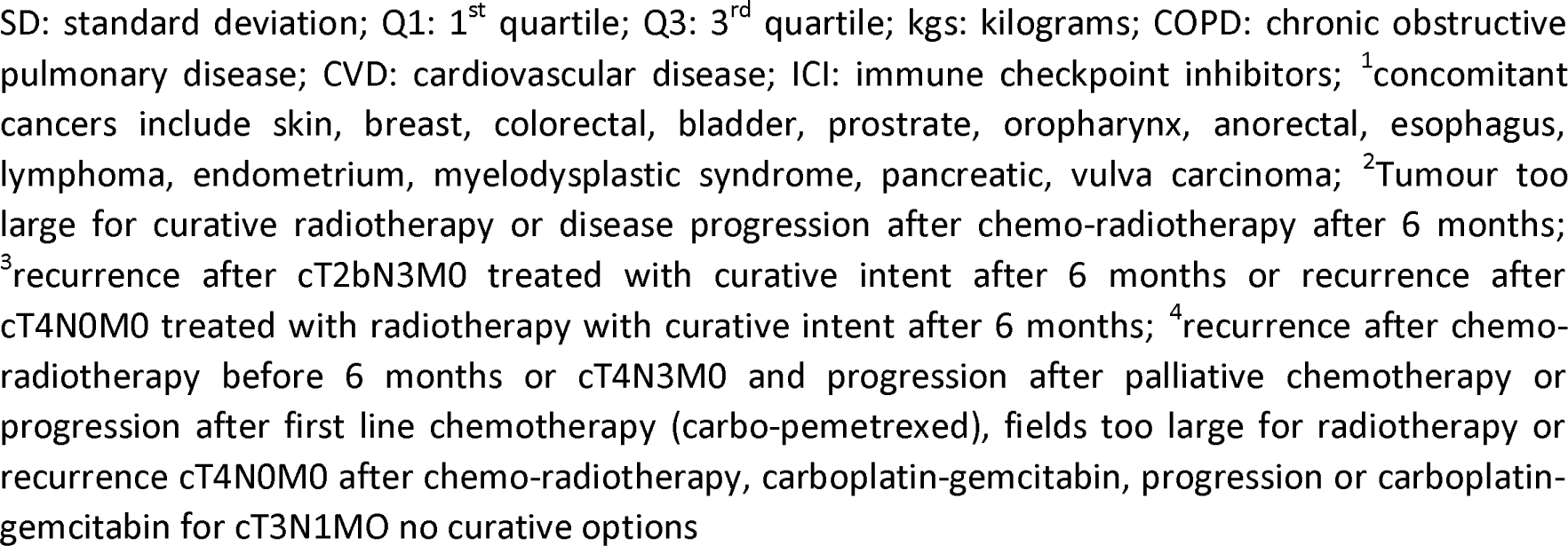
Baseline clinical and demographic characteristics of patients with lung cancer by treatment group.

| Variable | Overall | Treatment groups |  |  |  |
| --- | --- | --- | --- | --- | --- |
|  |  | Group 1:<br>chemo-radiotherapy<br>followed by<br>Durvalumab | Group 2:<br>immuno-monotherapy | Group 3:<br>immuno-chemotherapy | Group 4:<br>immunotherapy |
|  |  | 1<sup>st</sup> line | 1<sup>st</sup> line | 1<sup>st</sup> line | 2<sup>nd</sup>/further line |
|  |  | Curative intent | Palliative intent | Palliative intent | Palliative intent |
| Sample size (n,%) | 418 | 37 (8.9) | 70 (16.7) | 49 (11.7) | 262 (62.7) |
| Females (n,%) | 161 (38.5) | 13 (35.1) | 25 (35.7) | 14 (28.6) | 109 (41.6) |
| Age at ICI treatment (years) |  |  |  |  |  |
| Mean (SD) | 65.12 (9.03) | 63.70 (9.52) | 65.07 (9.78) | 67.47 (8.11) | 64.90 (8.89) |
| Median (Q1,Q3) | 66 (59,71) | 65 (61,71) | 66 (59,73) | 70 (62,73) | 66 (58,71) |
| Weight (kg) |  |  |  |  |  |
| Mean (SD) | 78.78 (15.96) | 81.26 (15.51) | 81.32 (18.76) | 82.52 (17.78) | 77.13 (14.73) |
| Median (Q1,Q3) | 77 (67,90) | 81 (71,93) | 80 (69,94) | 82 (73,93) | 75 (67,87) |
| Missing (n,%) | 11 (3) | 0 (0) | 4 (6) | 4 (8) | 3 (1) |
| ECOG performance status (n,%) |  |  |  |  |  |
| no limit to normal activity: 0 | 182 (43.5) | 26 (70.3) | 26 (37.1) | 20 (40.8) | 110 (42.0) |
| ambulatory, but restricted in strenuous activity: 1 | 204 (48.8) | 11 (29.7) | 35 (50.0) | 25 (51.0) | 133 (50.8) |
| able to perform self-care, active >50% of daytime: 2 | 22 (5.3) | 0 (0.0) | 7 (10.0) | 2 (4.1) | 13 (5.0) |
| only partly able to perform self-care, resting >50% of daytime: 3 | 4 (1.0) | 0 (0.0) | 1 (1.4) | 0 (0.0) | 3 (1.1) |
| Missing | 6 (1.4) | 0 (0.0) | 1 (1.4) | 2 (4.1) | 3 (1.1) |
| Diabetes (n,%) | 67 (16.0) | 8 (21.6) | 13 (18.6) | 11 (22.4) | 35 (13.4) |
| Hypertension (n,%) | 182 (43.5) | 20 (54.1) | 32 (45.7) | 26 (53.1) | 104 (39.7) |
| COPD (n,%) | 119 (28.5) | 11 (29.7) | 19 (27.1) | 12 (24.5) | 77 (29.4) |
| Rheumatological conditions (n,%) | 37 (8.9) | 5 (13.5) | 11 (15.7) | 3 (6.1) | 18 (6.9) |
| History of CVD (n,%) | 158 (37.8) | 18 (48.6) | 27 (38.6) | 24 (49.0) | 89 (34.0) |
| Concomitant cancer<sup>1</sup> (n,%) | 34 (8.1) | 1 (2.7) | 6 (8.6) | 6 (12.2) | 21 (8.0) |
| Education (n,%) |  |  |  |  |  |
| Low | 243 (58.1) | 21 (56.8) | 39 (55.7) | 28 (57.1) | 155 (59.2) |
| Medium | 71 (17.0) | 8 (21.6) | 12 (17.1) | 8 (16.3) | 43 (16.4) |
| High | 75 (17.9) | 7 (18.9) | 11 (15.7) | 12 (24.5) | 45 (17.2) |
| Missing | 29 (6.9) | 1 (2.7) | 8 (11.4) | 1 (2.0) | 19 (7.3) |
| Number of months since lung cancer diagnosis |  |  |  |  |  |
| Mean (SD) | 6.64 (8.20) | 4.43 (2.29) | 2.69 (4.94) | 1.39 (3.04) | 8.99 (9.11) |
| Median (Q1,Q3) | 4 (1,10) | 4 (3,5) | 2 (1,2) | 1 (0,2) | 7 (2,13) |
| Lung cancer stage (n,%) |  |  |  |  |  |
| 3 | 49 (11.7) | 36 (97.3) | 4<sup>2</sup> (5.7) | 2<sup>3</sup> (4.1) | 7<sup>4</sup> (2.7) |
| 4 | 369 (88.3) | 1 (2.7) | 66 (94.3) | 47 (95.9) | 255 (97.3) |

SD: standard deviation; Q1: 1<sup>st</sup> quartile; Q3: 3<sup>rd</sup> quartile; kgs: kilograms; COPD: chronic obstructive pulmonary disease; CVD: cardiovascular disease; ICI: immune checkpoint inhibitors; <sup>1</sup>concomitant cancers include skin, breast, colorectal, bladder, prostate, oropharynx, anorectal, esophagus, lymphoma, endometrium, myelodysplastic syndrome, pancreatic, vulva carcinoma; <sup>2</sup>Tumour too large for curative radiotherapy or disease progression after chemo-radiotherapy after 6 months; <sup>3</sup>recurrence after cT2bN3M0 treated with curative intent after 6 months or recurrence after cT4N0M0 treated with radiotherapy with curative intent after 6 months; <sup>4</sup>recurrence after chemo-radiotherapy before 6 months or cT4N3M0 and progression after palliative chemotherapy or progression after first line chemotherapy (carbo-pemetrexed), fields too large for radiotherapy or recurrence cT4N0M0 after chemo-radiotherapy, carboplatin-gemcitabin, progression or carboplatin-gemcitabin for cT3N1M0 no curative options

Covariables with missing values were ECOG PS (1.4%), weight (3.0%) and education (6.9%), which led to an exclusion of 40 (9.5%) patients in the complete-case analysis (Appendix 4). The results from the sensitivity analysis performed using multiple imputation of missing covariates, were similar to the complete-case analysis (see below).

Any clinically relevant changes in QoL over time is defined according to the evidence-based guidelines for interpreting changes in EORTC QLQ-C30 gQoL scores as no clinically significant change ([−5 to 5] points), small ([−10 to −5] / [5 to 8] points), medium ([−16 to −10] / [>8] points), or large ([< - 16] / not evaluable) deterioration/improvement^29^. These guidelines were developed by combining expert opinions and meta-analysis results from studies reporting QoL data using the EORTC QLQ-C30. While large, medium, and small differences were defined as those with unequivocal, probable, and subtle clinical relevance, respectively, trivial differences were defined as those likely to lack clinical relevance. Overall, there was no clinically significant change in predicted gQoL scores over 18 months of all lung cancer patients treated with ICI, conditional on their survival (Figure A.17). However, there were significant differences in these trajectories by treatment groups (Figure 2A). Patients with first-line immuno-monotherapy with palliative intent (group 2) had a small improvement ([5 to 8] points) in their gQoL score within the first six months of immunotherapy, and no clinically significant change ([−5 to 5] points) thereafter. Patients with first-line chemo-radiotherapy followed by Durvalumab with curative intent (group 1), had no clinically significant change in their gQoL over 18 months. Patients receiving first-line chemo-immunotherapy with a palliative intent (group 3) had a small improvement ([5 to 8] points) in their gQoL over 18 months. Group 4 patients with second/further-line immunotherapy with palliative intent had no clinically significant change in their gQoL over 18 months after immunotherapy (Figures 2A and A.13). Variation in predicted mean gQoL trajectories by ECOG PS and comorbidities are shown in Figures 2B-D. Patients with ‘fully active’ PS at baseline had higher gQoL over time compared to those who were ambulatory, but restricted in strenuous activity (PS=1). Patients with CVD or concomitant cancer had a lower gQoL compared to those who did not have these conditions respectively. Difference in predicted mean gQoL trajectories by other covariates were not clinically significant and hence not presented in this paper.

**Figure 2:**
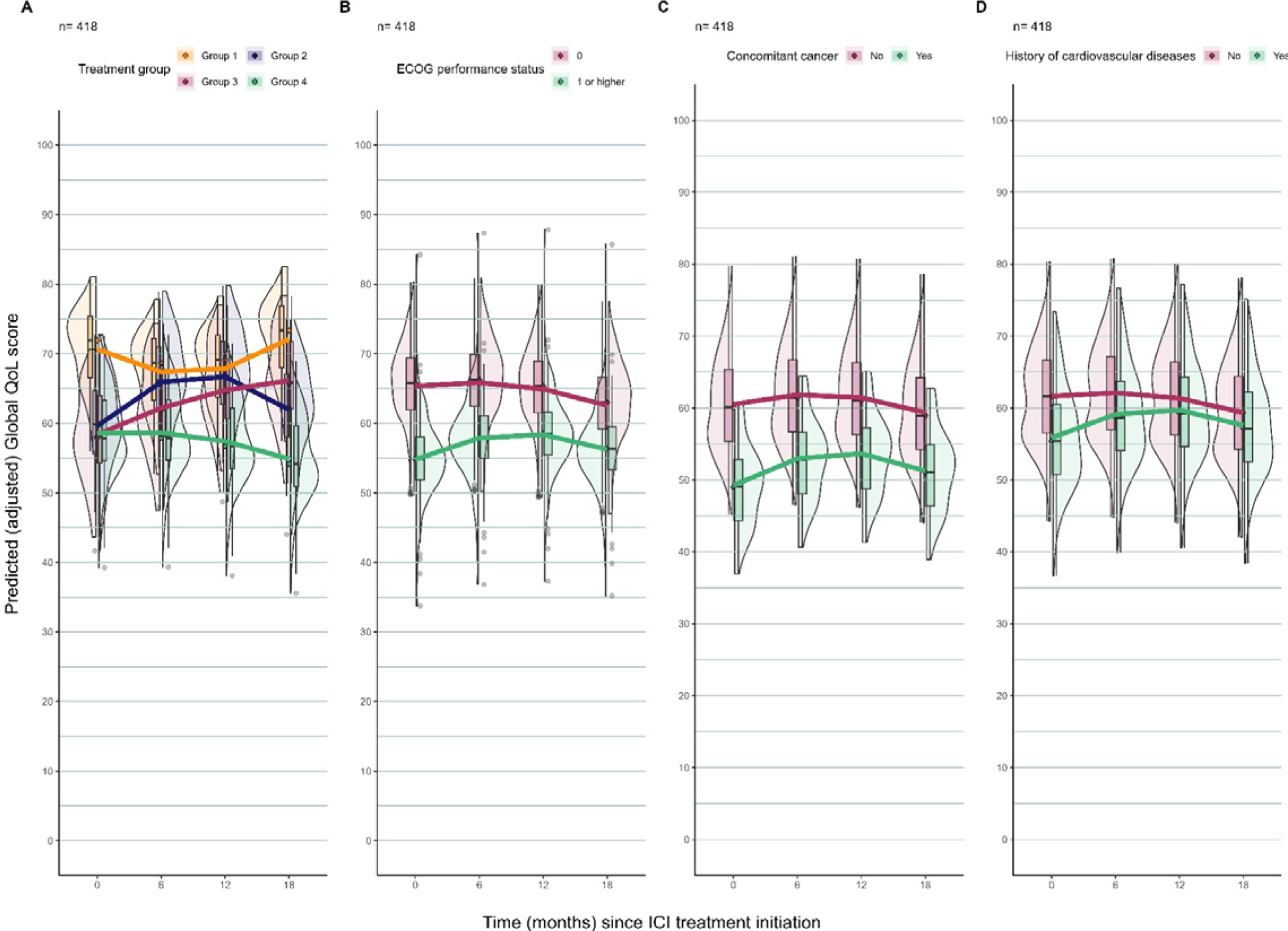
Predicted trajectory of gQoL scores of patients using joint model with multiple imputation accounting for the competing risk of death by A. treatment group, B. ECOG PS, C. Concomitant cancer, D. History of CVD. Violin plots summarise the subject-level predicted (adjusted) gQoL score and are superimposed with a curve showing the population-level predicted (adjusted) mean gQoL score for each sub-group of patients in the full cohort. Group-wise trajectories of predicted gQoL are presented here only for those groups which were significantly different. Group 1: first line chemo-radiotherapy followed by Durvalumab with curative intent; Group 2: first line immuno-monotherapy with palliative intent; Group 3: first line immune-chemotherapy with palliative intent; Group 4: second/further line immunotherapy with palliative intent; Patients with ECOG PS = 0 at baseline are those who are fully active; Patients with ECOG PS ≥ 1 at baseline are those who are restricted I physical activity.

First line immuno-monotherapy (versus second/further line immunotherapy) was associated with 6% higher gQoL over time (95% CI=[0.1%, 11%]). However, first line chemo-radiotherapy followed by Durvalumab, and first line immuno-chemotherapy (versus second/further line immunotherapy) were not associated with lower or higher gQoL over time. Having a history of CVD (posterior mean difference (PM)=-5%, 95% CI=[−10%, −0.1%]) or concomitant cancer at baseline (PM =-9%, 95% CI=[−17%, −2%]) were associated with 5% and 9% lower gQoL over time respectively. Being ambulatory, but restricted in strenuous activity (PS=1) at baseline (versus fully active, PS=0) was associated with a 9% lower gQoL over time (PM_PS=1_=-9%, 95% CI=[−13%, −5%]) after adjustment on other covariates and accounting for the competing risk of death. Other comorbidities (diabetes, hypertension, COPD and rheumatological conditions), tumour stage, number of months since tumour diagnosis, age, sex, weight, and education were not associated with the gQoL over time (Table A.3).

The results from sensitivity analysis after imputing education under a MNAR mechanism (as ‘low’ or ‘high’) were similar to models fitted with multiple imputation (Figures A.17-A.19) giving reassurance that our results were robust to departure from the MAR assumption assumed for multiple imputation.

The predicted mean trajectory of the five functional scores of lung cancer patients from baseline to up to 18 months from the joint model with multiple imputation is shown in Figure 3. While the physical ([−10 to −5] points) and role ([−14 to −7] points) functioning had a small deterioration over 18 months, changes in emotional ([−3 to 6] points), social ([−6 to −3] points), and cognitive ([−1 to 3] points) functioning were not clinically significant, conditional on patients’ survival.

**Figure 3:**
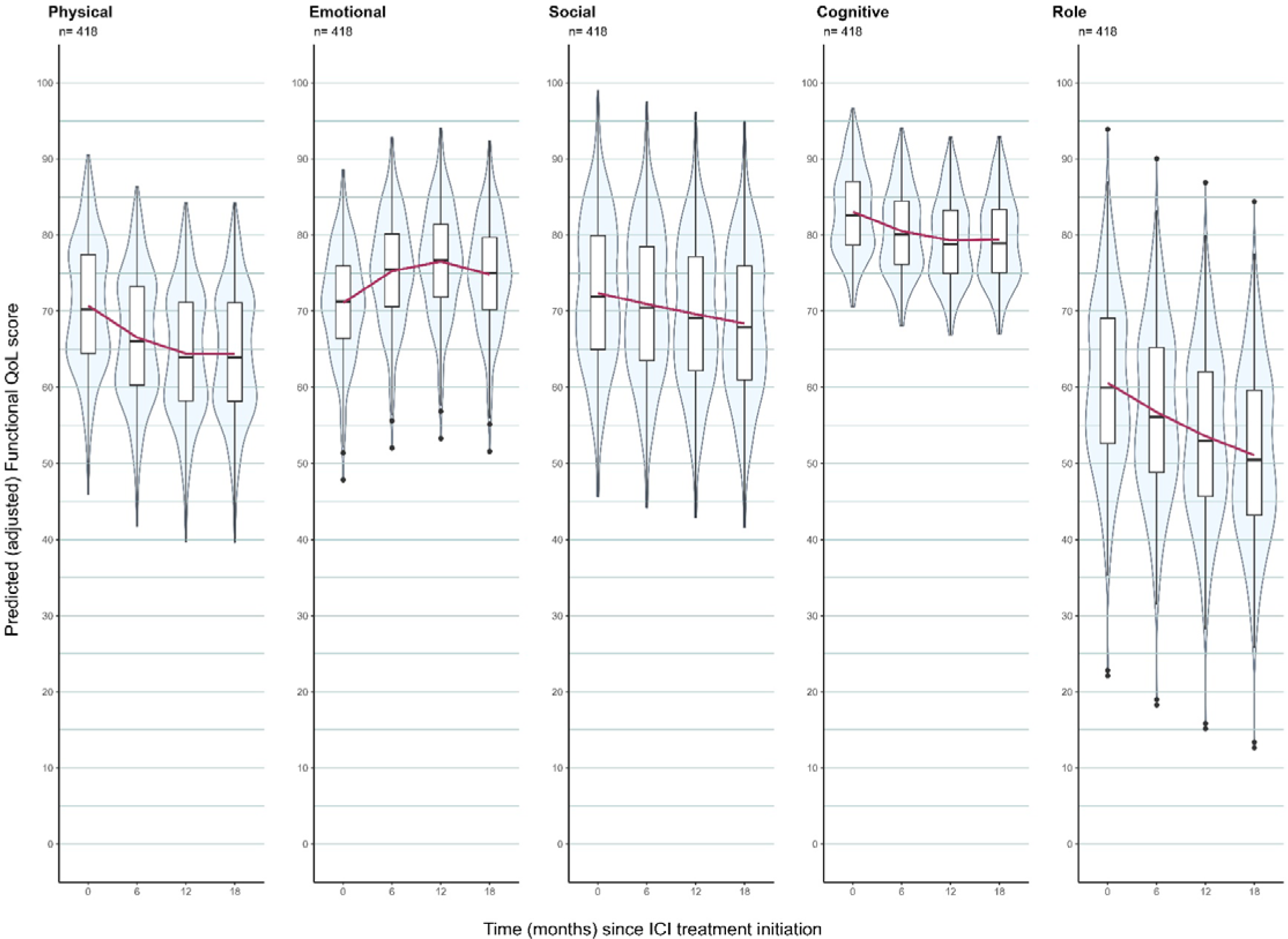
Predicted trajectory of functional scale scores from a Joint model with multiple imputation to account for missing data for the full cohort. Violin plots summarise the subject-level predicted (adjusted) functional QoL score and are superimposed with a curve showing the population-level predicted (adjusted) functional score for the full cohort.

## Discussion

QoL indicators measured by patient-reported outcomes are pivotal in assessing the effect of cancer treatments on the QoL of patients with advanced stage lung cancer. Such patients may experience a change in their QoL over time due to their prognosis and treatment-related side-effects. Hence, research and clinical practice are ascending towards optimising patients’ QoL^30^. The changes in QoL of lung cancer patients following ICI treatment has only received limited and recent attention in research^31–33^. In this longitudinal cohort study, we aimed at studying the changes in gQoL of patients with advanced stage lung cancer who were treated with ICIs and other cancer treatments at a tertiary cancer hospital in the Netherlands. Using joint modelling which accounted for survival time and key confounders, we showed that there were differences in the trajectories of gQoL of patients with different treatment regimens based on its intensity, line, and intent. As expected, patients who had treatment was with curative intent (group 1) had a higher baseline gQoL than those with palliative treatments (groups 2-4), reflecting less extensive disease (mainly stage 3) and/or better PS (generally = 0). However, because the treatment intensity was maximal (chemo-radiotherapy followed by Durvalumab) among those patients (group 1), they tended to have more side-effects from the treatment associated with a transient decrease in their gQoL over 18 months. Overall, the presence or absence of chemotherapy in the treatment seemed to be associated with transient deterioration (group 1) or improvement (group 2) of the gQoL, respectively. This agrees with previous studies which have shown ICIs to be associated with higher QoL and longer time to clinical deterioration compared to chemotherapy alone in different types of solid tumours^31 32^. However, in agreement with other studies^33 34^, we found a small improvement in the gQoL of patients treated by Pembrolizumab, a PD-1 inhibitor, concurrently with chemotherapy (group 3), even if this improvement was initially slower than in the immuno-monotherapy group (group 2).

QoL of patients with lung cancer treated with ICIs and pre-existing CVD is a complex and important consideration. Previous studies have shown that pre-existing CVD among lung cancer patients treated with immunotherapy is associated with poorer overall survival^35^. Our analysis showed that lung cancer patients with a history of CVD had a lower gQoL over time (versus those with no CVD), which is a strong prognostic factor for survival in NSCLC patients^36^. Since the incidence of clinically significant symptoms, impacting QoL, is greater among patients with advanced stage lung cancer and poor PS^37^, we also observed a lower gQoL over time among those with relatively poor PS (≥1) in our cohort. Patients with concomitant cancer within one year of ICI treatment initiation had a lower gQoL over time in our analysis, however, we did not find any evidence from literature studying the impact of concomitant cancer on QoL. There was no association between age at ICI initiation and changes in gQoL in our study, which also supports the results of a recent study based on OncoLifeS data-biobank^14^ of lung cancer patients treated with ICIs. A study on older patients with advanced lung cancer treated with systemic therapy reported a deterioration in physical functioning over a period of six months^38^, which agrees with our results. There were no studies yet showing the changes in other EORTC functional scales among lung cancer patients treated with ICIs.

### Strengths and limitations

Our analyses were based on a large set of real-world data where patients were followed longitudinally over a period of 18 months after their ICI treatment. Linkage of these data with clinical records allowed us to adjust for important covariates. We have studied QoL using the EORTC QLQ-C30, a tool widely used in oncological research^39^, which helped us to compare our results with similar studies. With repeated measurements on QoL, we developed a model which predicted the QoL trajectory of patients with advanced stage lung cancer, from the start of their ICI treatment to up to 18 months. Exploration of the association between missingness and covariates, outcomes and treatment suggested some evidence of an outcome-dependent MAR mechanism. We addressed this issue of missing data in the covariates and performed a sensitivity analysis using multiple imputation under a Bayesian framework. We also checked the robustness of our results by imputing missing observations in ‘education’ under a MNAR mechanism and compared them with the results obtained using multiple imputation. Since the results were similar, we believe our results were robust to some departure of the MAR assumption postulated for multiple imputation.

Specifically for lung cancer, the problem of incomplete questionnaires from dropouts or missing data from deceased patients during the observation period was inevitable^40^. Hence, we could not perform our analysis on the full two-year period, and instead had to restrict it to 18 months. There was a possibility for selection bias as patients with higher QoL were more likely to participate in the study than patients with lower QoL^41^. If patient’s health deteriorated after treatment, their participation would have drastically reduced. There was an absence of a control group of patients who did not have ICIs but only received other anti-cancer treatments such as chemotherapy. This did not allow us to compare the evolution of gQoL over time between those who had only immunotherapy versus those who had only chemotherapy. We could not assess the interaction effect of treatment groups and PS on QoL due to small numbers in these sub-groups.

### Conclusion and recommendation for clinical practice

Our findings suggest that the differences in trajectories of gQoL over time among patients with varying treatment regimens based on drug combination as well as line and intent of treatment may help guide clinicians and patients of potential benefits and impairments of treatments on QoL. This may further help identify patients who need additional care during their treatment for lung cancer at advanced stages. Since decision making is largely driven by randomised trials, which do not provide a full picture because of their restricted inclusion criteria, hence, assessing the benefits of treatments through QoL measurements based on observational data is crucial.

## Acronyms

QoL – quality of life

gQoL – global quality of life

ICI – immune checkpoint inhibitors IRAEs - immune-related adverse events

## Data availability

The OncoLifeS data-biobank follows the requirements of the European General Data Protection Regulation for scientific research. Data from the OncoLifeS database can be accessed via a protected workspace after approval of the steering committee. Interested researchers may contact the OncoLifeS initiative directly (http://www.oncolifes.nl) to inquire about access to the data.

## Acknowledgement

We thank all participants enrolled in OncoLifeS database who consented to use their medical data for this study. We also thank the developers and managers of OncoLifeS data-biobank for sharing their data with us.

## Author contributions

Conceptualisation: Ananya Malhotra, Clemence Leyrat, Manuela Quaresma and Bernard Rachet. Methodology: Ananya Malhotra, Clemence Leyrat, Manuela Quaresma and Bernard Rachet. Formal analysis: Ananya Malhotra. Data curation: Ananya Malhotra, Erick Suazo Zepeda. Writing – original draft: Ananya Malhotra. Writing – review and editing: all co-authors. Supervision: Manuela Quaresma, Clemence Leyrat and Bernard Rachet.

## Ethics declaration

This study was approved by the London School of Hygiene & Tropical Medicine (LSHTM) Research Ethics Committee. OncoLifeS has been approved by the medical ethics committee of the UMCG (UMCG METC approval 2010/109) and has been ISO certified (9001:2008 Healthcare). It was registered in the Dutch Trial Register under the number: NL7839. Participants of the OncoLifeS data biobank are included only after a written informed consent form.

