## Appendices for "Changes in Global Quality of Life after treatment with immune checkpoint inhibitors in patients with advanced stage lung cancer in the Netherlands – a 2015-2021 cohort study"

**Appendix 1**

|  | Variable / Field Name | Field Label | Field Attributes (Field Type, Validation, Choices, Calculations, etc.) |
| --- | --- | --- | --- |
| Patient and tumor characteristics | | | |
| 1 | [record_id] | Record ID (OncoLifeS Number) | text |
| 2 | [date_weight_height] | Date of weight and height assessment  **(Always measured at baseline, the closest date to the start of the immunotherapy)** | text (date_dmy) |
| 3 | [height] | Height (meters)  **(Always measured at baseline, the closest date to the start of the immunotherapy)** | text (number) |
| 4 | [weight] | Weight (kilograms)  **(Always measured at baseline, the closest date to the start of the immunotherapy)** | text (number) |
| 5 | [bmi] | BMI  **(Always measured at baseline, the closest date to the start of the immunotherapy)** | Calculation: [weight]/([height]*[height]) |
| 6 | [date_performance_status] | Date of Performance Status assessment  **(Always measured at baseline, the closest date to the start of the immunotherapy)** | text (date_dmy) |
| 7 | [performance_status] | Performance status  **(Always measured at baseline, the closest date to the start of the immunotherapy)** | radio 1 \| 0; 2 \| 1; 3 \| 2; 4 \| 3; 5 \| 4; 6 \| 5 |
| 8 | [diabetes] | History of type II diabetes  **(Always measured at baseline, the closest date to the start of the immunotherapy)** | 1 \| Yes; 2 \| No; 3 \| Unknown |
| 9 | [treatment_dm] | Treatment diabetes | text |
| 10 | [hypertension] | History of hypertension  Defined as either: -Measured in the hospital with the value of 140mmHg for systolic and/or 90mmHg for diastolic blood pressure -Use of hypertensive drugs | 1 \| Yes; 2 \| No; 3 \| Unknown |
| 11 | [treatment_hyper] | Treatment for hypertension | text |
| 12 | [copd] | History of chronic obstructive pulmonary disease  **(Always measured at baseline, the closest date to the start of the immunotherapy)** | 1 \| Yes; 2 \| No; 3 \| Unknown |
| 13 | [treatment_copd] | Treatment for COPD | text |
| 14 | [rheumatological_conditions] | History of rheumatological conditions (e.g., Lupus, rheumatoid arthritis, Sjogren's Syndrome)  **(Always measured at baseline, the closest date to the start of the immunotherapy)** | 1 \| Yes; 2 \| No; 3 \| Unknown |
| 15 | [treatment_reuma] | Treatment for rheumatological disease | text |
| 16 | [dementia] | Presence of dementia  **(Always measured at baseline, the closest date to the start of the immunotherapy)** | 1 \| Yes; 2 \| No; 3 \| Unknown |
| 17 | [treatment_dement] | Treatment for dementia | text |
| 18 | [history_of_cardiovascular] | History of cardiovascular disease (cerebrovascular disease, myocardial infarction)  **(Always measured at baseline, the closest date to the start of the immunotherapy)** | 1 \| Yes; 2 \| No; 3 \| Unknown |
| 19 | [treatment_cardiovasc] | Treatment for cardiovascular disease | text |
| 20 | [date_diagnosis] | Date of diagnosis | text (date_dmy) |
| 21 | [date_clinical_staging] | Date of staging (Clinical staging) | text (date_dmy) |
| 22 | [tnm_classification_t_clin] | TNM classification, T (Clinical T) | 1 \| Tx; 2 \| T0; 3 \| Tis; 4 \| T1; 5 \| T1a; 6 \| T1b; 7 \| T1c; 8 \| T2; 9 \| T2a; 10 \| T3; 11 \| T4; 12 \| Unknown; 13 \| T2b |
| 23 | [tnm_classification_n_clin] | TNM classification, N (Clinical N) | 1 \| N1; 2 \| N2; 3 \| N3; 5 \| N0; 4 \| Unknown |
| 24 | [tnm_classification_m_clin] | TNM classification, M (Clinical M) | 1 \| No metastasis; 2 \| M1; 3 \| M1a; 4 \| M1b; 5 \| M1c; 6 \| Unknown |
| 25 | [tnm_version_clin] | TNM classification version (Clinical) | 1 \| 8th version; 2 \| 7th version; 3 \| Unknown |
| 26 | [stage_tnm_clin] | Stage (Clinical) | 1 \| I; 2 \| IA; 3 \| IB; 4 \| II; 5 \| IIA; 6 \| IIB; 7 \| III; 8 \| IIIA; 9 \| IIIB; 10 \| IIIC; 11 \| IV; 12 \| IVA; 13 \| IVB |
| 27 | [date_path_staging] | Date of staging (Pathological staging)  **(Only when reported)** | text (date_dmy) |
| 28 | [tnm_classification_t_path] | TNM classification, T (Pathology T) | 1 \| Tx; 2 \| T0; 3 \| Tis; 4 \| T1; 5 \| T1a; 6 \| T1b; 7 \| T1c; 8 \| T2; 9 \| T2a; 10 \| T3; 11 \| T4; 12 \| Unknown |
| 29 | [tnm_classification_n_path] | TNM classification, N (Pathology N) | 1 \| N1; 2 \| N2; 3 \| N3; 5 \| N0; 4 \| Unknown |
| 30 | [tnm_classification_m_path] | TNM classification, M (Pathology M) | 1 \| No metastasis; 2 \| M1; 3 \| M1a; 4 \| M1b; 5 \| M1c; 6 \| Unknown |
| 31 | [tnm_version_path] | TNM classification version (Pathology) | 1 \| 8th version; 2 \| 7th version; 3 \| Unknown |
| 32 | [stage_tnm_path] | Stage (Pathology) | 1 \| I; 2 \| IA; 3 \| IB; 4 \| II; 5 \| IIA; 6 \| IIB; 7 \| III; 8 \| IIIA; 9 \| IIIB; 10 \| IIIC; 11 \| IV; 12 \| IVA; 13 \| IVB |
| 33 | [histological_type] | Histological type | 1 \| Non-small-cell lung cancer; 2 \| Small cell lung cancer; 3 \| Non-small-cell lung cancer (Adenocarcinoma); 4 \| Non-small-cell lung cancer (Squamous cell carcinoma); 5 \| Non-small-cell lung cancer (Large cell carcinoma); 6 \| Other |
| 34 | [other_histo_type] | Describe other histological type | text |
| 35 | [pd1_expression] | PD-L1 expression | 1 \| < 1; 2 \| =>1 - < 50%; 3 \| < 50%; 4 \| Unknown/not tested |
| 36 | [mutations] | Mutations | 1 \| Not tested; 2 \| Tested, no mutation; 3 \| Squamous cell carcinoma, no mutation; 4 \| AK; 5 \| AKT1; 6 \| ALK; 7 \| BRAF; 8 \| C-KIT; 9 \| CMET; 10 \| EGFR; 11 \| ESR1; 12 \| FGFR1; 13 \| GNA; 14 \| GNAQ; 15 \| GNAS; 16 \| H3F3A; 17 \| H3F3B; 18 \| HER2; 19 \| HRAS; 20 \| IDH-1; 21 \| IDH-2; 22 \| IDH-3; 23 \| JAK2; 24 \| KRAS; 25 \| MAP2K1; 26 \| MEK; 27 \| MET; 28 \| NRAS; 29 \| NRG1; 30 \| NTRK1; 31 \| NTRK2; 32 \| NTRK3; 33 \| PDGFRA; 34 \| PIK3CA; 35 \| POLE1; 36 \| PTEN; 37 \| RET; 38 \| ROS; 39 \| ROS1; 40 \| T790M; 41 \| TP53; 42 \| Other |
| 37 | [concominant_cancer] | Concomitant cancer  **(Only other cancers diagnosed within one year before the start of the immunotherapy)** | 1 \| Yes; 2 \| No |
| 38 | [type_of_concomitant_cancer] | Type of concomitant cancer | text |
| 39 | [date_diag_concomit_can] | Date of diagnosis, concomitant cancer | text (date_dmy) |
| 40 | [patient_and_tumor_character istics_complete] | Complete? | 0 \| Incomplete; 1 \| Unverified; 2 \| Complete |
| Laboratory findings | | | |
| 41 | [date_immuno_lab] | Date of the test  **(The most recent study up to two weeks prior to the start of immunotherapy)** | text (date_dmy) |
| 42 | [leukocytes] | Leukocytes (10^9 cells/l)  **(The most recent study up to two weeks prior to the start of immunotherapy)** | text (number) |
| 43 | [neutrophiles] | Neutrophiles (10^9 cells/l)  **(The most recent study up to two weeks prior to the start of immunotherapy)** | text (number) |
| 44 | [lymphocytes] | Lymphocytes (10^9 cells/l)  **(The most recent study up to two weeks prior to the start of immunotherapy)** | text (number) |
| 45 | [eosinophils] | Eosinophils (10^9 cells/l)  **(The most recent study up to two weeks prior to the start of immunotherapy)** | text (number) |
| 46 | [date_crp] | Date of the test  **(The most recent study up to two weeks prior to the start of immunotherapy)** | text (date_dmy) |
| 47 | [crp] | CRP (mg/l)  **(The most recent study up to two weeks prior to the start of immunotherapy)** | text (number) |
| 48 | [date_of_liver_function] | Date of liver function assessment  **(The most recent study up to two weeks prior to the start of immunotherapy)** | text (date_dmy) |
| 49 | [alt] | ALAT (U/L)  **(The most recent study up to two weeks prior to the start of immunotherapy)** | text (number) |
| 50 | [alp] | ALP (U/L)  **(The most recent study up to two weeks prior to the start of immunotherapy)** | text (number) |
| 51 | [ast] | ASAT (U/L)  **(The most recent study up to two weeks prior to the start of immunotherapy)** | text (number) |
| 52 | [ggt] | GGT (U/L)  **(The most recent study up to two weeks prior to the start of immunotherapy)** | text (number) |
| 53 | [date_of_kidney_function] | Date of kidney function assessment  **(The most recent study up to two weeks prior to the start of immunotherapy)** | text (date_dmy) |
| 54 | [creatinine] | Creatinine (umol/L)  **(The most recent study up to two weeks prior to the start of immunotherapy)** | text (number) |
| 55 | [date_of_thyroid_function] | Date of thyroid function assessment  **(The most recent study up to two weeks prior to the start of immunotherapy)** | text (date_dmy) |
| 56 | [tsh] | TSH (mU/L)  **(The most recent study up to two weeks prior to the start of immunotherapy)** | text (number) |
| 57 | [date_cortisol] | Date of cortisol assessment  **(The most recent study up to two weeks prior to the start of immunotherapy)** | text (date_dmy) |
| 58 | [cortisol] | Cortisol (nmol/L)  **(The most recent study up to two weeks prior to the start of immunotherapy)** | text (number) |
| 59 | [ldh_date] | LDH test date  **(The most recent study up to two weeks prior to the start of immunotherapy)** | text (date_dmy) |
| 60 | [ldh_level] | LDH (U/L)  **(The most recent study up to two weeks prior to the start of immunotherapy)** | text (number) |
| 61 | [albumine_date] | Albumin test date  **(The most recent study up to two weeks prior to the start of immunotherapy)** | text (date_dmy) |
| 62 | [albumine_level] | Albumin level (g/L) | text (number) |
| 63 | [faecal_pcr_test_performed] | Fecal viral PCR test performed?  **(Performed prior or during the immunotherapy treatment)** | 1 \| Yes; 0 \| No |
| 64 | [date_fecal_viral_test] | Date fecal viral test  **(Performed prior or during the immunotherapy treatment)** | text (date_dmy) |
| 65 | [fecal_viral_pcr_result] | Fecal viral PCR result  **(Performed prior or during the immunotherapy treatment)** | text |
| 66 | [faecal_culture_performed] | Fecal culture performed?  **(Performed prior or during the immunotherapy treatment)** | 1 \| Yes; 0 \| No |
| 67 | [date_faecal_bacter_study] | Date of fecal bacteriology study  **(Performed prior or during the immunotherapy treatment)** | text (date_dmy) |
| 68 | [faecal_culture_result] | Fecal culture result (Bacterial study)  **(Performed prior or during the immunotherapy treatment)** | text |
| 69 | [anti_nuclear_antibodies] | Anti-nuclear antibodies  **(Performed prior to the start of Immunotherapy, no limit of time)** | 1 \| Positive; 2 \| Negative; 3 \| Unknown/not tested |
| 70 | [ana_date] | Anti-nuclear antibodies test date  **(Performed prior to the start of Immunotherapy, no limit of time)** | text (date_dmy) |
| 71 | [anca] | Antineutrophil cytoplasmic antibody (ANCA)  **(Performed prior to the start of Immunotherapy, no limit of time)** | 1 \| Positive; 2 \| Negative; 3 \| Unknown/not tested |
| 72 | [anca_date] | ANCA test date  **(Performed prior to the start of Immunotherapy, no limit of time)** | text (date_dmy) |
| 73 | [rheumatoid_factor] | Rheumatoid factor (IU/ml)  **(Performed prior to the start of Immunotherapy, no limit of time)** | text (number) |
| 74 | [rheumatoid_factor_date] | Rheumatoid factor test date  **(Performed prior to the start of Immunotherapy, no limit of time)** | text (date_dmy) |
| 75 | [laboratory_findings_complete] | Complete? | 0 \| Incomplete; 1 \| Unverified; 2 \| Complete |
| Immunotherapy | | | |
| 76 | [monotherapy] | Monotherapy | 1 \| Yes; 0 \| No |
| 77 | [treatment_line] | Treatment line | 1 \| First line; 2 \| Second line; 3 \| Third or higher line |
| 78 | [start_date_immunotherapy_1] | Start date immunotherapy | text (date_dmy) |
| 79 | [stop_date_immunotherapy_1] | Stop date immunotherapy | text (date_dmy) |
| 80 | [num_cycles_immunother] | Number of immunotherapy cycles | text (number) |
| 81 | [purpose_of_the_treatment] | Purpose of the treatment  **(As stated in the clinical record)** | 1 \| Adjuvant; 2 \| Neoadjuvant; 3 \| Curative; 4 \| Palliative |
| 82 | [immunotherapy_agent_1] | Name of immunotherapy agent 1 | text |
| 83 | [dose_immunotherapy] | Dose of immunotherapy agent | text (number) |
| 84 | [dose_units] | Dose units | 1 \| µg; 2 \| mg; 3 \| mg/kg; 4 \| USP; 5 \| mg/m2; 6 \| AUC |
| 85 | [immuno_modific] | Did the original treatment regime suffered modifications (i.e. double doses due to COVID-19 pandemic)? | 1 \| Yes; 0 \| No |
| 86 | [immunotherapy_ongoing] | Is the immunotherapy treatment still ongoing (5-11-2021) ? | 1 \| Yes; 0 \| No |
| 87 | [immuno_modif_descri] | Describe the treatment regime modifications | text |
| 88 | [immunotherapy_agent_2] | Name of immunotherapy agent 2 | text |
| 89 | [cycles_immunot_2] | Number of cycles immunotherapy agent 2 | text (number) |
| 90 | [dose_immunotherapy_2] | Dose of immunotherapy agent 2 | text (number) |
| 91 | [dose_units_2] | Dose units 2 | 1 \| µg; 2 \| mg; 3 \| mg/kg; 4 \| USP; 5 \| mg/m2; 6 \| AUC |
| 92 | [immunotherapy_agent_3] | Name of immunotherapy agent 3 | text |
| 93 | [cycles_immunot_3] | Number of cycles immunotherapy agent 3 | text (number) |
| 94 | [dose_immunotherapy_3] | Dose of immunotherapy agent 3 | text (number) |
| 95 | [dose_units_3] | Dose units 3 | 1 \| µg; 2 \| mg; 3 \| mg/kg; 4 \| USP; 5 \| mg/m2; 6 \| AUC |
| 96 | [immunotherapy_agent_4] | Name of immunotherapy agent 4 | text |
| 97 | [cycles_immunot_4] | Number of cycles immunotherapy agent 4 | text (number) |
| 98 | [dose_immunotherapy_4] | Dose of immunotherapy agent 4 | text (number) |
| 99 | [dose_units_4] | Dose units 4 | 1 \| µg; 2 \| mg; 3 \| mg/kg; 4 \| USP; 5 \| mg/m2; 6 \| AUC |
| 100 | [chemotherapy_agent_1] | Name of chemotherapy agent 1 | text |
| 101 | [cycles_chemo_1] | Number of cycles chemotherapy agent 1 | text (number) |
| 102 | [dose_chemotherapy_1] | Dose of chemotherapy agent 1 | text (number) |
| 103 | [dose_units_chemo_1] | Dose units chemotherapy 1 | 1 \| µg; 2 \| mg; 4 \| mg/kg; 3 \| USP; 5 \| mg/m2; 6 \| AUC |
| 104 | [chemotherapy_agent_2] | Name of chemotherapy agent 2 | text |
| 105 | [cycles_chemo_2] | Number of cycles chemotherapy agent 2 | text (number) |
| 106 | [dose_chemotherapy_2] | Dose of chemotherapy agent 2 | text (number) |
| 107 | [dose_units_chemo_2] | Dose units chemotherapy 2 | 1 \| µg; 2 \| mg; 4 \| mg/kg; 3 \| USP; 5 \| mg/m2; 6 \| AUC |
| 108 | [immuno_early_stop] | Did the immunotherapy stop early? | 1 \| Yes; 2 \| No |
| 109 | [reason_early_stop] | Reason for early stop | 1 \| Disease progression; 2 \| Immune related adverse events; 4 \| Transfer to another hospital; 3 \| Other |
| 110 | [reason_early_stop_other] | describe Other | text |
| 111 | [treatment_response] | Treatment response (RECIST)-Complete response, disappearance of all lesion and pathologic lymph nodes-Partial response, ≥30% decrease in the sum of the longest diameters, no new lesions, no progression of non-target lesions-Stable disease, no partial response or progressive disease met- Progressive disease, ≥20% increase in the sum of the longest diameters compared to the smallest sum of the longest diameters, or progression of the non-target lesions or new lesions | 1 \| Complete response; 2 \| Partial response; 3 \| Stable disease; 4 \| Progressive disease |
| 112 | [date_treatment_resp] | Date of evaluation of response to treatment | text (date_dmy) |
| 113 | [best_overall_response] | Best overall treatment response. The best overall response is the best response recorded from the start of the study treatment until the end of treatment | 1 \| Complete response; 2 \| Partial response; 3 \| Stable disease; 4 \| Progressive disease |
| 114 | [date_of_best_overall_treat] | Date of Best Overall treatment response assessment | text (date_dmy) |
| 115 | [treatment_response_after_6w] | Treatment response after 6 weeks from immunotherapy stop date | 1 \| Complete response; 2 \| Partial response; 3 \| Stable disease; 4 \| Progressive disease |
| 116 | [date_treatment_response_6w] | Date of treatment response assessment after 6 weeks | text (date_dmy) |
| 117 | [treatment_response_after_3] | Treatment response after 3 months from immunotherapy stop date | 1 \| Complete response; 2 \| Partial response; 3 \| Stable disease; 4 \| Progressive disease |
| 118 | [date_treatment_response_3] | Date of treatment response assessment after 3 months | text (date_dmy) |
| 119 | [treatment_response_after_6] | Treatment response after 6 months from immunotherapy stop date | 1 \| Complete response; 2 \| Partial response; 3 \| Stable disease; 4 \| Progressive disease |
| 120 | [date_treatment_response_6] | Date of treatment response assessment after 6 months | text (date_dmy) |
| 121 | [treatment_response_12] | Treatment response after 12 months from immunotherapy stop date | 1 \| Complete response; 2 \| Partial response; 3 \| Stable disease; 4 \| Progressive disease |
| 122 | [date_treatment_response_12] | Date of treatment response assessment after 12 months | text (date_dmy) |
| 123 | [immunotherapy_complete] | Complete? | 0 \| Incomplete; 1 \| Unverified; 2 \| Complete |
| Immune-related adverse events (irAEs) | | | |
| 124 | [iraes] | Did the patient develop immune related adverse events with this treatment? | 1 \| Yes; 0 \| No |
| 125 | [date_irae_reported] | Date irAE reported | text (date_dmy) |
| 126 | [descripition_irae] | Description of immune related adverse event 1 (According to CTCAE) | text |
| 127 | [irae_treatment] | Did the immune related adverse events require withdrawal of treatment or any extra treatment? | 1 \| No extra treatment; 2 \| Treatment withdrawal; 3 \| Extra treatment; 4 \| Extra treatment and treatment withdrawal |
| 128 | [irae_treat_description] | Describe the irAE treatment | text |
| 129 | [start_date_irae_treatment] | Start date irAE treatment | text (date_dmy) |
| 130 | [stop_date_irae_treatment] | Stop date irAE treatment (or when corticosteroids < 5mg/day) | text (date_dmy) |
| 131 | [irae_treat_max_dose] | Maximum dose irAE treatment | text (number) |
| 132 | [irae_treat_max_dose_2] | Dose Units | 1 \| µg; 2 \| mg; 3 \| mg/kg; 4 \| USP; 5 \| mg/m2; 6 \| AUC |
| 133 | [manteinance_dose] | Corticosteroid maintenance dose (enter 0 when treatment stopped) | text Field Annotation: Include frequency and unit |
| 134 | [ae_not_irae] | Is this adverse event likely to be NOT related to the immunotherapy, but to chemotherapy agents? | 1 \| Yes; 0 \| No |
| 135 | [date_of_iraes_collection] | Date of irAEs collection | text (date_dmy) |
| 136 | [iraes_complete] | Complete? | 0 \| Incomplete; 1 \| Unverified; 2 \| Complete |
| Other treatments | | | |
| 137 | [radiotherapy] | Was the patient treated with radiotherapy?  **(Prior or during the Immunotherapy treatment)** | 1 \| Yes; 0 \| No |
| 138 | [radiotherapy_start_date] | Start date of most recent radiotherapy regimen  **(Prior or during the Immunotherapy treatment)** | text (date_dmy) |
| 139 | [radiotherapy_start_date_2] | Stop date of most recent radiotherapy regimen  **(Prior or during the Immunotherapy treatment)** | text (date_dmy) |
| 140 | [radiotherapy_target] | Radiotherapy target | 1 \| Lung; 2 \| Brain; 3 \| Bones; 4 \| Other |
| 141 | [radiotherapy_target_other] | Radiotherapy target, other | text |
| 142 | [surgery] | Did the patient undergo a major surgery?  Major surgery defined as: A invasive procedure in which an extensive resection is performed, a body cavity is entered, organs are removed, or normal anatomy is altered. In general, if a mesenchymal barrier is opened (pleural cavity, peritoneum, meninges)  **(Prior or during the Immunotherapy treatment)** | 1 \| Yes; 0 \| No |
| 143 | [surgery_type] | Type of most recent major surgery  **(Prior or during the Immunotherapy treatment)** | text |
| 144 | [surgery_date] | Date of most recent major surgery  **(Prior or during the Immunotherapy treatment)** | text (date_dmy) |
| 145 | [chemotherapy] | Was the patient treated with chemotherapy?  **(Prior Immunotherapy treatment)** | 1 \| Yes; 0 \| No |
| 146 | [chemotherapy_start_date] | Start date of most recent chemotherapy regimen  **(Prior Immunotherapy treatment)** | text (date_dmy) |
| 147 | [chemotherapy_stop_date] | Stop date of most recent chemotherapy regimen  **(Prior Immunotherapy treatment)** | text (date_dmy) |
| 148 | [targeted_therapy] | Was the patient treated with targeted therapy? | 1 \| Yes; 0 \| No |
| 149 | [start_date_targeted] | Start date targeted therapy | text (date_dmy) |
| 150 | [stop_date_targeted] | stop date targeted therapy | text (date_dmy) |
| 151 | [other_treatments_complete] | Complete? | 0 \| Incomplete; 1 \| Unverified; 2 \| Complete |
| Quality of life questionnaires | | | |
| 152 | [date_baseline_qol] | Date of filling baseline QoL questionnaire. | text (date_dmy) |
| 153 | [date_6month_qol] | Date of filling the 6 months follow up QoL questionnaire. | text (date_dmy) |
| 154 | [date_12month_qol] | Date of filling the 12 months follow up QoL questionnaire. | text (date_dmy) |
| 155 | [date_18month_qol] | Date of filling the 18 months follow up QoL questionnaire. | text (date_dmy) |
| 156 | [date_24month_qol] | Date of filling the 24 months follow up QoL questionnaire. | text (date_dmy) |
| 157 | [quality_of_life_questionnaires_complete] | Complete? | dropdown 0 \| Incomplete; 1 \| Unverified; 2 \| Complete |
| Treatment after Immunotherapy stop | | | |
| 158 | [immunotherapy_after_immuno] | Did the patient continue with Immunotherapy, within three years of first starting immunotherapy treatment? | 1 \| Yes; 0 \| No; 2 \| Unknown (moved to a different hospital to continue their treatment) |
| 159 | [start_immuno_after_immuno] | Start date immunotherapy | text (date_dmy) |
| 160 | [stop_immuno_after_immuno] | Stop date immunotherapy | text (date_dmy) |
| 161 | [stop_immuno_after_immuno_2] | Stop date immunotherapy (2nd treatment) | text (date_dmy) |
| 162 | [start_immuno_after_immuno_2] | Start date immunotherapy (2nd treatment) | text (date_dmy) |
| 163 | [start_immuno_after_immuno_3] | Start date immunotherapy (3rd treatment) | text (date_dmy) |
| 164 | [stop_immuno_after_immuno_3] | Stop date immunotherapy (3rd treatment) | text (date_dmy) |
| 165 | [chemo_after_immuno] | Did the patient continue with chemotherapy, within three years of first starting immunotherapy treatment? | 1 \| Yes; 0 \| No; 2 \| Unknown (moved to a different hospital to continue their treatment) |
| 166 | [start_chemo_after_immuno] | Start date Chemotherapy | text (date_dmy) |
| 167 | [stop_chemo_after_immuno] | Stop date Chemotherapy | text (date_dmy) |
| 168 | [start_chemo_after_immuno_2] | Start date Chemotherapy (2nd treatment) | text (date_dmy) |
| 169 | [stop_chemo_after_immuno_2] | Stop date Chemotherapy (2nd treatment) | text (date_dmy) |
| 170 | [start_chemo_after_immuno_3] | Start date Chemotherapy(3rd treatment) | text (date_dmy) |
| 171 | [stop_chemo_after_immuno_3] | Stop date Chemotherapy(3rd treatment) | text (date_dmy) |
| 172 | [targetted_after_immuno] | Did the patient continue with targeted therapy, within three years of first starting immunotherapy treatment? | 1 \| Yes; 0 \| No; 2 \| Unknown (moved to a different hospital to continue their treatment) |
| 173 | [start_target_after_immuno] | Start date targeted | text (date_dmy) |
| 174 | [stop_target_after_immuno] | Stop date targeted | text (date_dmy) |
| 175 | [start_target_after_immu_2] | Start date targeted (2nd treatment) | text (date_dmy) |
| 176 | [stop_target_after_immuno_2] | Stop date targeted (2nd treatment) | text (date_dmy) |
| 177 | [start_target_after_immu_3] | Start date targeted (3rd treatment) | text (date_dmy) |
| 178 | [stop_target_after_immuno_3] | Stop date targeted(3rd treatment) | text (date_dmy) |
| 179 | [radio_after_immuno] | Did the patient continue with radiotherapy, within three years of first starting immunotherapy treatment? | 1 \| Yes; 0 \| No; 2 \| Unknown (moved to a different hospital to continue their treatment) |
| 180 | [start_radio_after_immuno] | Start date radiotherapy | text (date_dmy) |
| 181 | [stop_radio_after_immuno] | Stop date radiotherapy | text (date_dmy) |
| 182 | [start_radio_after_immuno_2] | Start date radiotherapy (2nd treatment) | text (date_dmy) |
| 183 | [stop_radio_after_immuno_2] | Stop date radiotherapy (2nd treatment) | text (date_dmy) |
| 184 | [start_radio_after_immuno_3] | Start date radiotherapy (3rd treatment) | text (date_dmy) |
| 185 | [stop_radio_after_immuno_3] | Stop date radiotherapy (3rd treatment) | text (date_dmy) |
| 186 | [surgery_after_immuno] | Did the patient had a surgery, within three years of first starting immunotherapy treatment? | 1 \| Yes; 0 \| No; 2 \| Unknown (moved to a different hospital to continue their treatment) |
| 187 | [date_surgery_after_immuno] | Date surgery | text (date_dmy) |
| 188 | [patient_died] | Has the patient died? | 1 \| Yes; 0 \| No |
| 189 | [date_of_death] | date of death | text (date_dmy) |
| 190 | [date_of_treatments_after_i] | Date of "Treatments after immunotherapy" data collection | text (date_dmy) |
| 191 | [treatments_after_immunotherapy_complete] | Complete? | 0 \| Incomplete; 1 \| Unverified; 2 \| Complete |

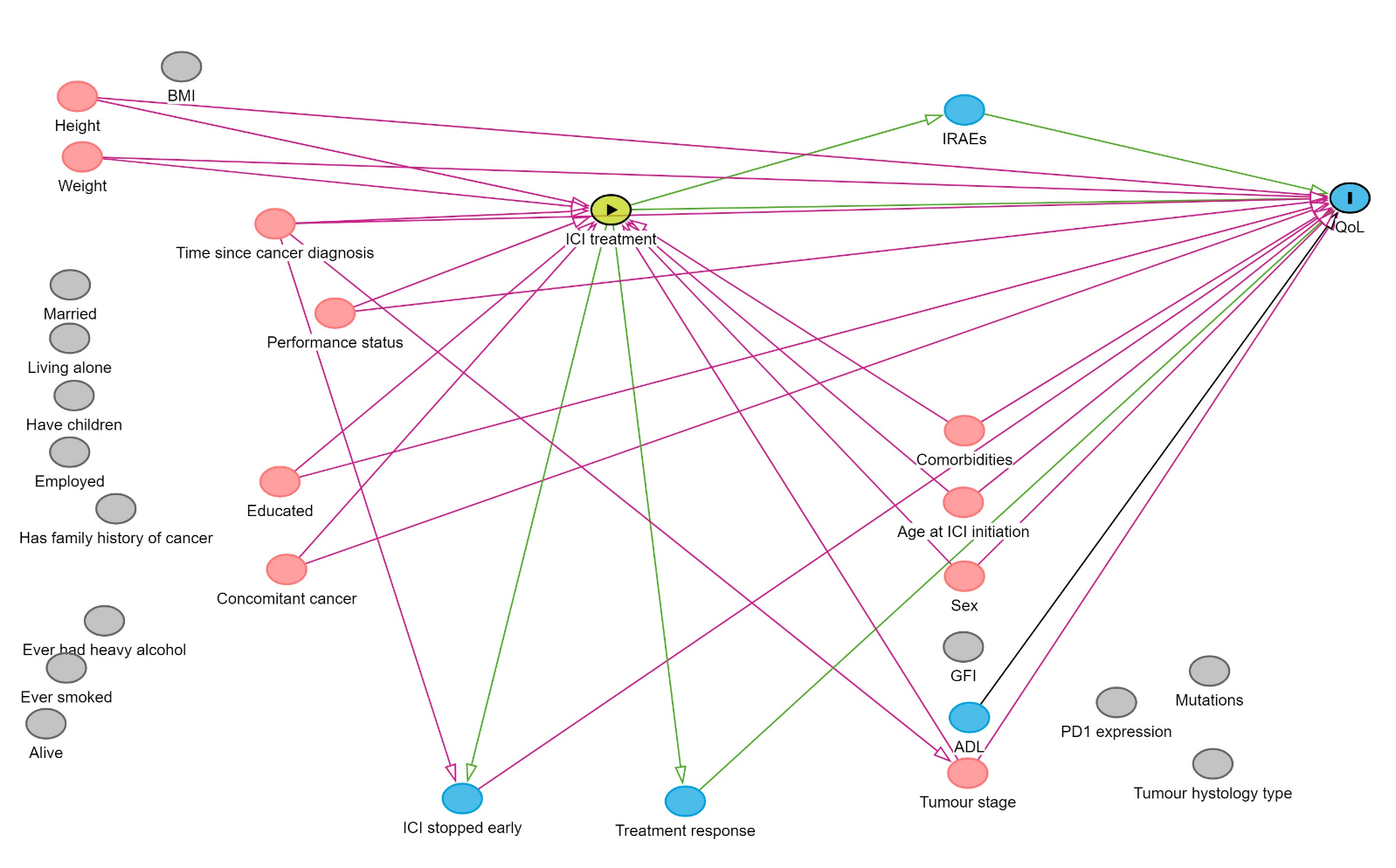
**Appendix 2**

**Figure A.1:** Directed Acyclic Graph where the exposure is ICI treatment and the outcome is QoL after ICI treatment. The variables in red are potential confounders and adjusted in the analysis for testing the association between ICI treatment and QoL. The variables in blue are mediators (as they lie in the causal pathway of the exposure and the outcome) which are not included in the analysis. The variables in grey are not included in the analysis because they are not associated with the exposure or the outcome.

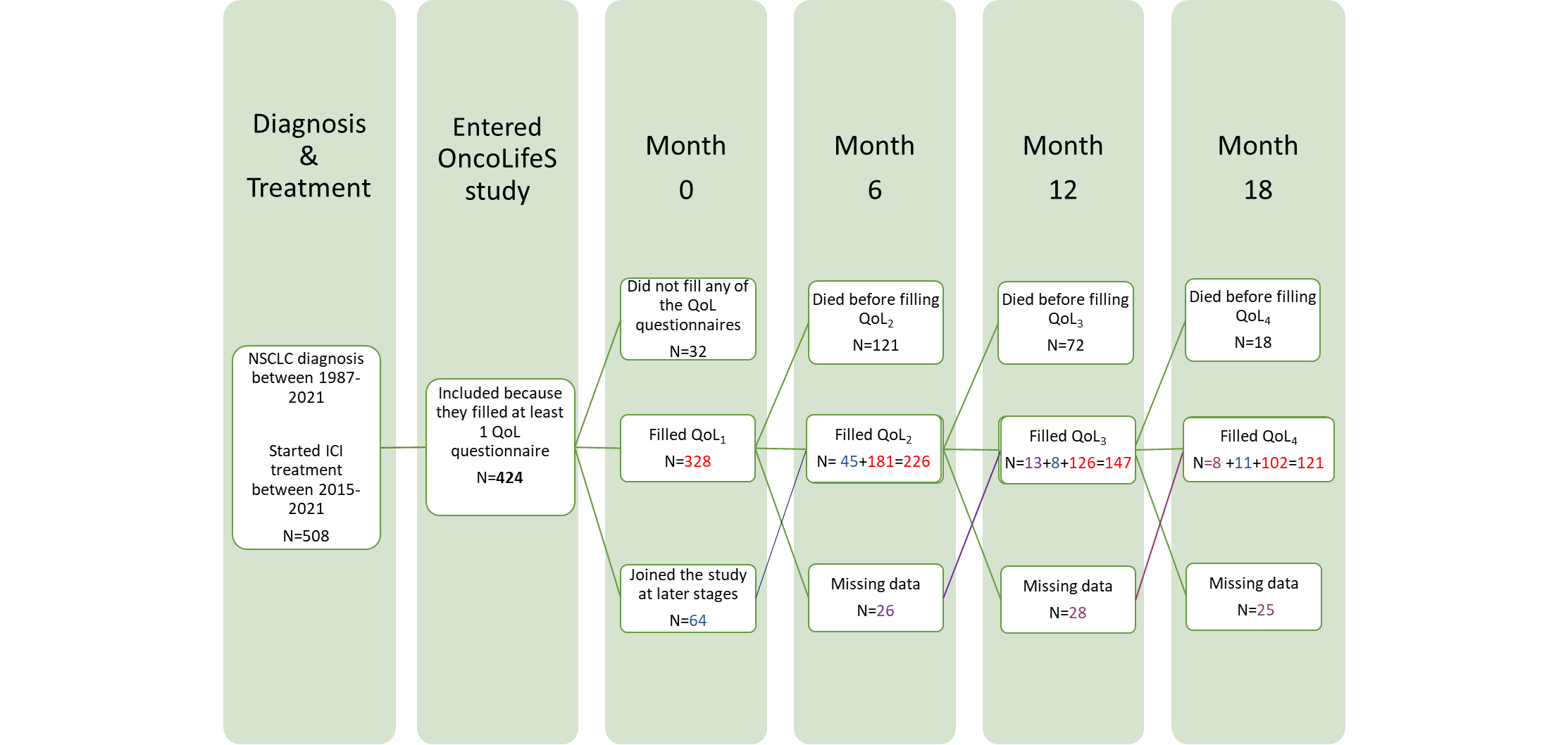

**Figure A.2:** Consort diagram showing the timeline of filling EORTC QLQ-C30 questionnaires. N is the number of patients (all stages) and the colours indicate the following:

- Number of patients who filled follow-up questionnaires given they filled all the previous questionnaires
- Number of patients who joined the study at later stages
- Number of patients who do not have a complete follow-up over 1.5 years because they missed their follow-up

**Appendix 3**

**Observed trajectory of functional scale scores**

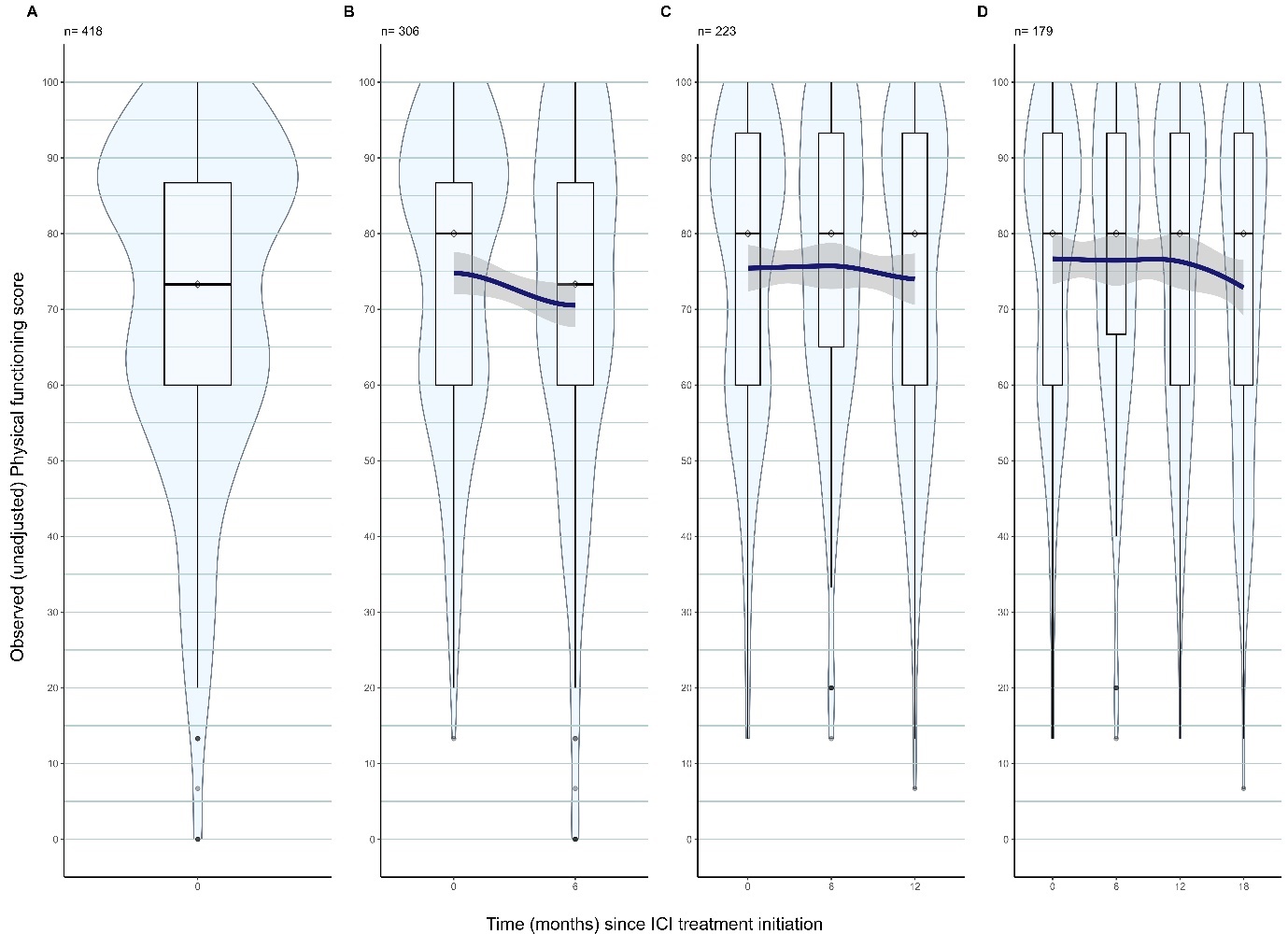

**Figure A.3:** Violin plots with superimposed smoothing curve (fitted using weighted least squares) describing the evolution of physical functioning scores among patients who survived after ICI treatment, where ‘n’ is the number of patients alive at baseline (panel A), up to 6 months (panel B), up to 12 months (panel C), and up to 18 months (panel D).

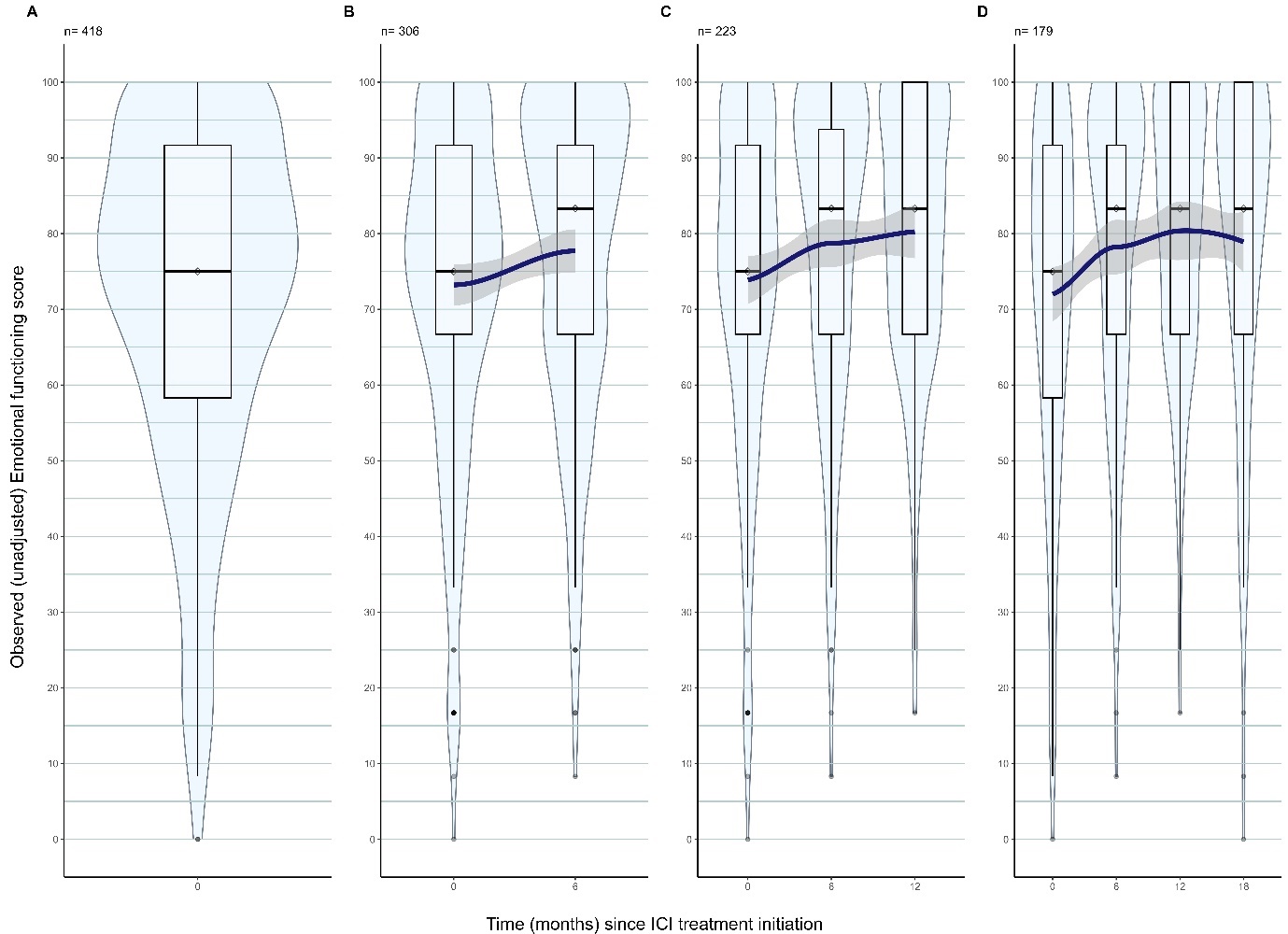

**Figure A.4:** Violin plots with superimposed smoothing curve (fitted using weighted least squares) describing the evolution of emotional functioning scores among patients who survived after ICI treatment, where ‘n’ is the number of patients alive at baseline (panel A), up to 6 months (panel B), up to 12 months (panel C), and up to 18 months (panel D).

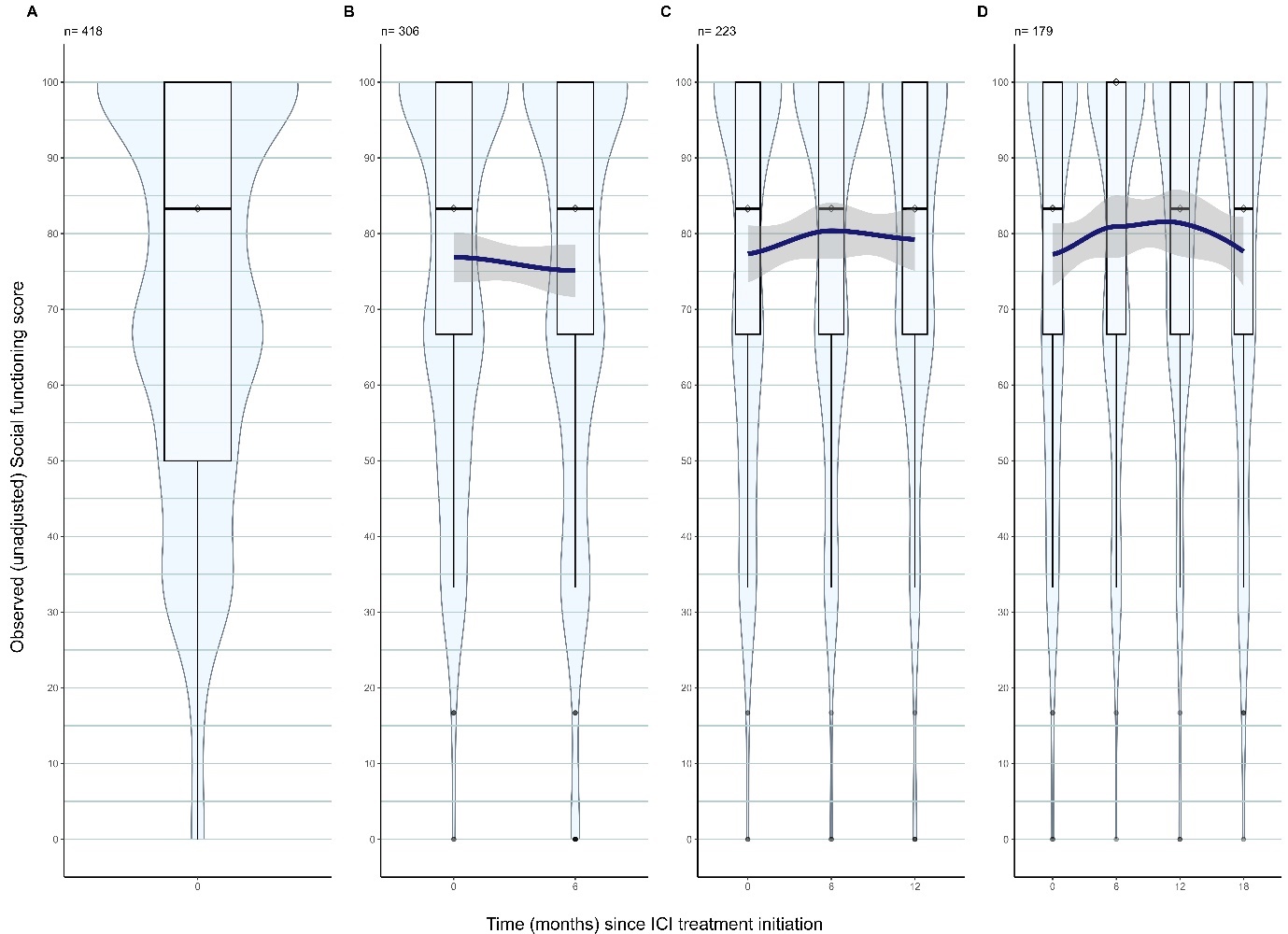

**Figure A.5:** Violin plots with superimposed smoothing curve (fitted using weighted least squares) describing the evolution of social functioning scores among patients who survived after ICI treatment, where ‘n’ is the number of patients alive at baseline (panel A), up to 6 months (panel B), up to 12 months (panel C), and up to 18 months (panel D).

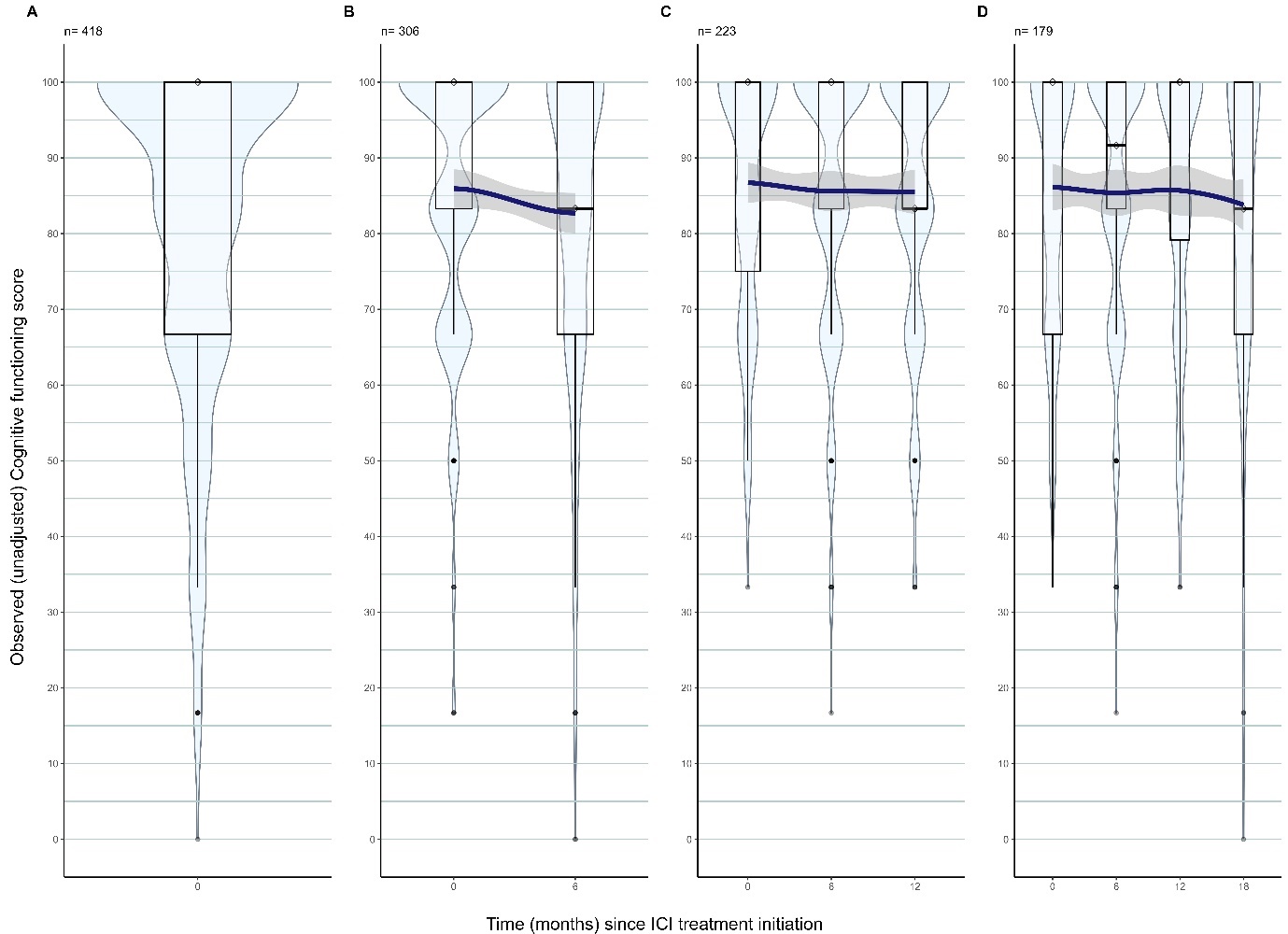

**Figure A.6:** Violin plots with superimposed smoothing curve (fitted using weighted least squares) describing the evolution of cognitive functioning scores among patients who survived after ICI treatment, where ‘n’ is the number of patients alive at baseline (panel A), up to 6 months (panel B), up to 12 months (panel C), and up to 18 months (panel D).

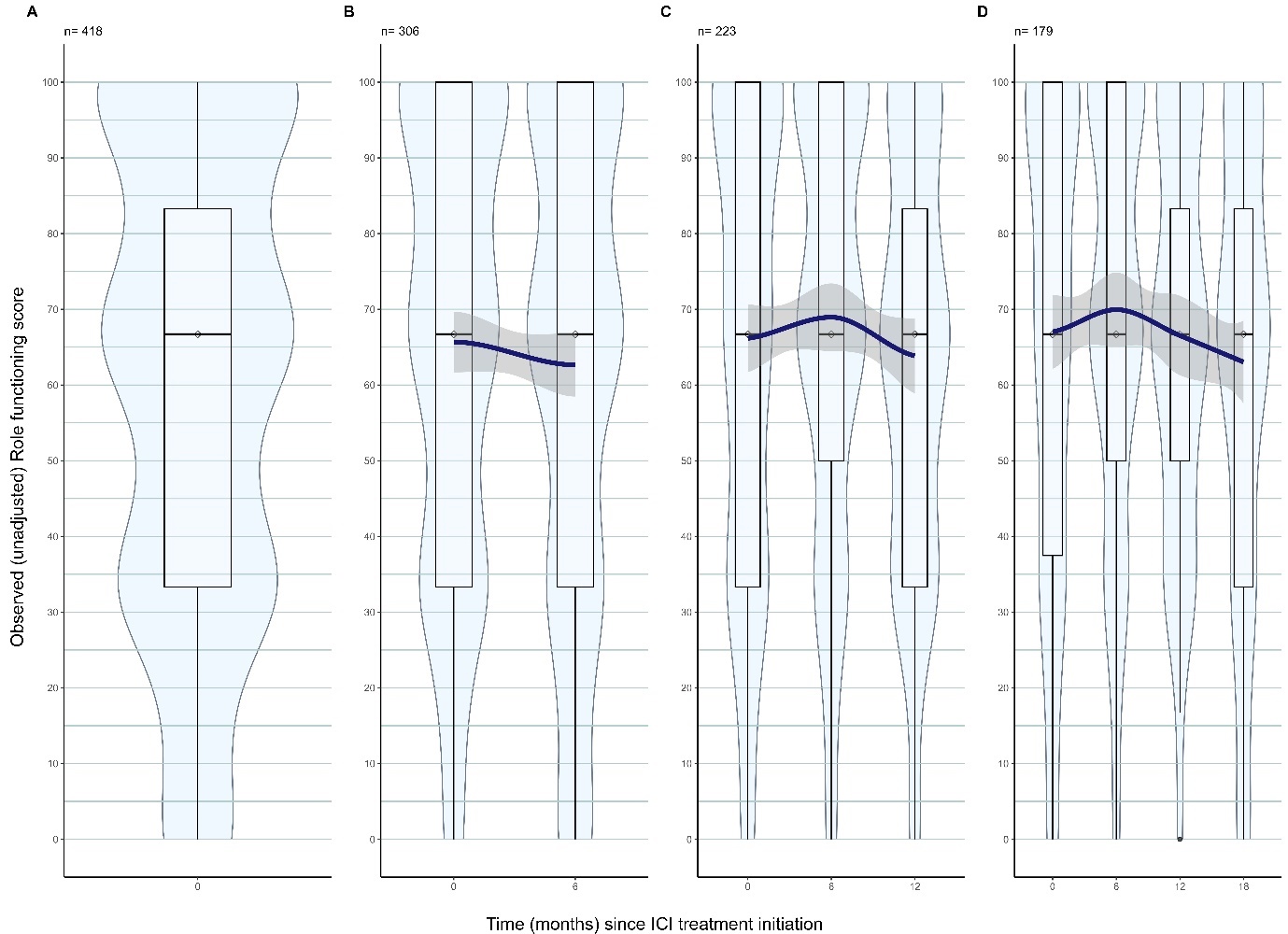

**Figure A.7:** Violin plots with superimposed smoothing curve (fitted using weighted least squares) describing the evolution of role functioning scores among patients who survived after ICI treatment, where ‘n’ is the number of patients alive at baseline (panel A), up to 6 months (panel B), up to 12 months (panel C), and up to 18 months (panel D).

**Appendix 4**

**Complete-case analysis**

**Model description**

The outcome here was the gQoL which was measured repeatedly over time, first at baseline and then every 6 months to up to 18 months. The exposure was a categorical variable of treatment groups, where group 1 consists of patients who had first line chemo-radiotherapy followed by durvalumab within 6 weeks to 3 months with a curative intent, group 2 consists of patients who had first line immuno-monotherapy with a palliative intent and no other prior anti-cancer treatments, group 3 consists of patients who had first line immune-chemotherapy with a palliative intent and group 4 consists of patients who had second or further line immunotherapy (either alone or in combination with chemotherapy) with a palliative intent. The covariables adjusted in the analysis were age, sex, weight, education, PS, comorbidities, concomitant cancer, tumour stage and number of years since cancer diagnosis. Observation time for QoL measurements and time-to-death since ICI treatment initiation were coded as continuous variables on the same scale (years). A quadratic term for time of QoL measurement (observation time) was added to reflect the shape of the (unadjusted) relationship between QoL score and observation time (Figure 1). To facilitate model convergence and interpretation, variables age and weight were standardised (centred at their mean). The gQoL score was scaled by dividing by 100 so it lies between 0 and 1.

Adjusted mean difference (AMD) and its 95% confidence interval (CI) was used to describe the measure of association between gQoL and covariates. Model summaries are presented in Table A.2 below. The predicted mean gQoL trajectory of lung cancer patients from baseline to up to 18 months from the joint model is shown in Figure A.8. To assess the differences between the trajectory of predicted mean gQoL by treatment groups, we included an additional interaction term between observation time of QoL and treatment group (Figure A.8). Similarly, an additional interaction term between observation time of QoL and comorbidities was included in the model where we assessed the differences between the trajectory of predicted mean gQoL by presence of comorbidities at baseline (Figures A.9-A.12).

The histogram of the random effects from the joint model was normally distributed. The joint model diagnostic plots including the plot of marginal standardised residuals versus fitted values, the plot of subject-specific residuals versus fitted values and the Q-Q plot of subject-specific residuals, showed no systematic trend.

**Results**

**Table A.2:** Model summary from linear mixed effect sub-model of the joint model fitted using *JM* package in R

| **Variable** |  | **95% CI of AMD** | |
| --- | --- | --- | --- |
|  | **AMD** | **2.50%** | **97.50%** |
| Intercept | 0.62 | 0.50 | 0.74 |
| Observation time for QoL | -0.03 | -0.10 | 0.05 |
| Observation time for QoL^2^ | 0.00 | -0.05 | 0.05 |
| Treatment group 1 ( vs group 4) | 0.12 | -0.01 | 0.25 |
| Treatment group 2 ( vs group 4) | 0.04 | -0.02 | 0.10 |
| Treatment group 3 ( vs group 4) | 0.04 | -0.03 | 0.11 |
| Weight | 0.02 | 0.00 | 0.05 |
| ECOG PS = 1 (vs PS = 0) | -0.11 | -0.15 | -0.07 |
| ECOG PS = 2 (vs PS = 0) | -0.12 | -0.22 | -0.02 |
| ECOG PS = 3 (vs PS = 0) | -0.07 | -0.30 | 0.15 |
| Diabetes (vs no) | -0.02 | -0.07 | 0.04 |
| Have hypertension (vs no) | 0.03 | -0.01 | 0.08 |
| Have COPD (vs no) | 0.01 | -0.03 | 0.06 |
| Have rheumatological conditions (vs no) | -0.02 | -0.10 | 0.05 |
| Have history of CVD (vs no) | -0.06 | -0.10 | -0.01 |
| Have concomitant cancer (vs no) | -0.10 | -0.18 | -0.02 |
| Age | -0.01 | -0.03 | 0.02 |
| Female sex | 0.02 | -0.03 | 0.07 |
| Stage 4 (vs Stage = 3) | 0.02 | -0.10 | 0.14 |
| Medium-level Education (vs Low-level) | -0.03 | -0.08 | 0.03 |
| High-level Education (vs Low-level) | 0.00 | -0.05 | 0.06 |
| Number of months since lung cancer diagnosis | 0.01 | -0.02 | 0.04 |

AMD: adjusted mean difference

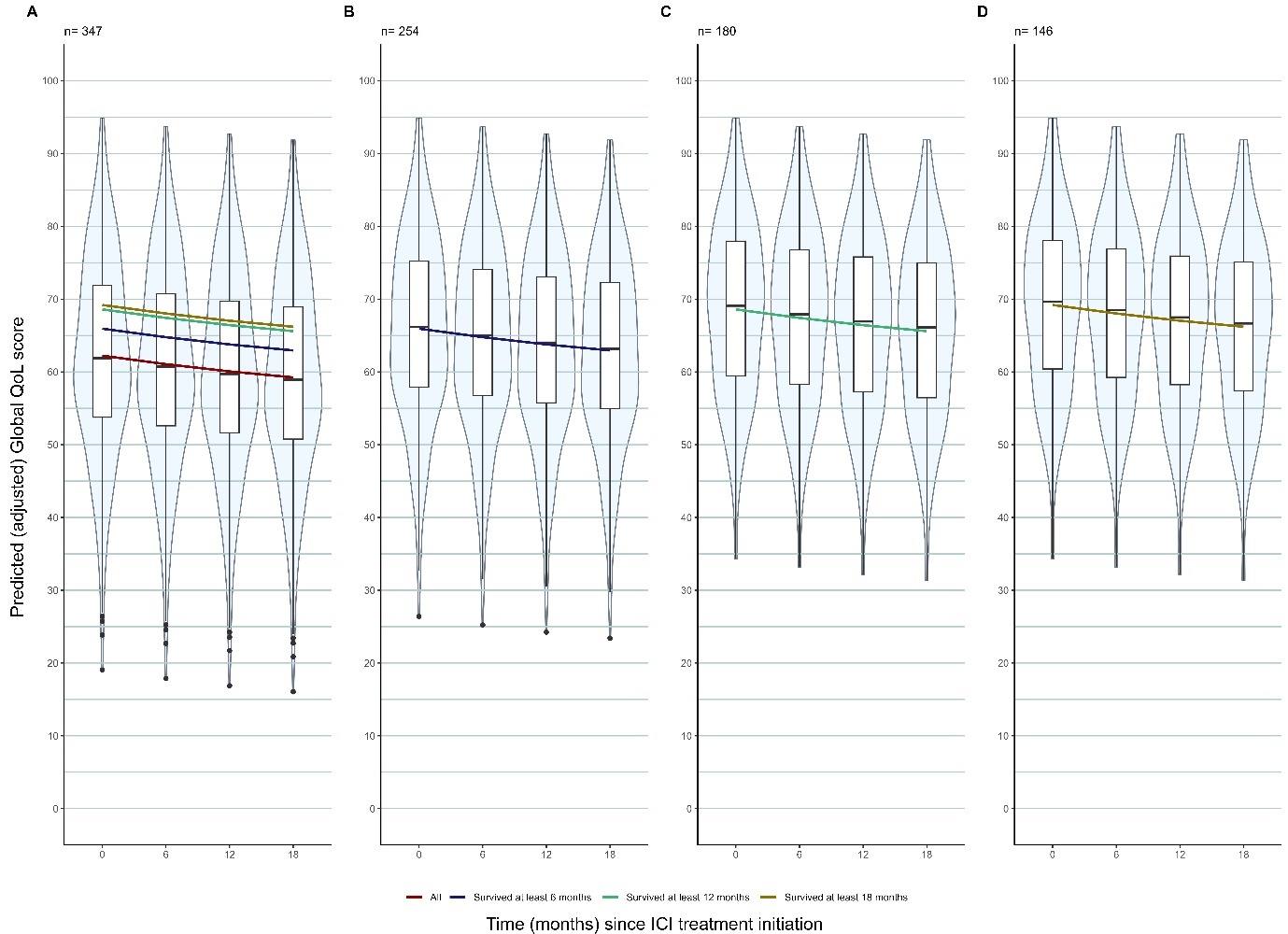

**Figure A.8**: Predicted trajectory of Global QoL scores from a Joint model with ‘observation time’ as a time-dependant continuous covariate plus a quadratic term for observation time and adjusted for confounders and competing risk of death using a Cox-PH model (complete case analysis). Violin plots of the subject-level predicted (adjusted) gQoL score are superimposed with a curve showing the population-level predicted (adjusted) gQoL score for A. the full cohort, B. those who survived at least 6 months, C. those who survived at least 12 months, D. those who survived at least 18 months. Panel A. also shows all other population level-predicted (adjusted) gQoL scores from panels B-D on a single graph depicting the relative difference in the trajectories among these cohorts.

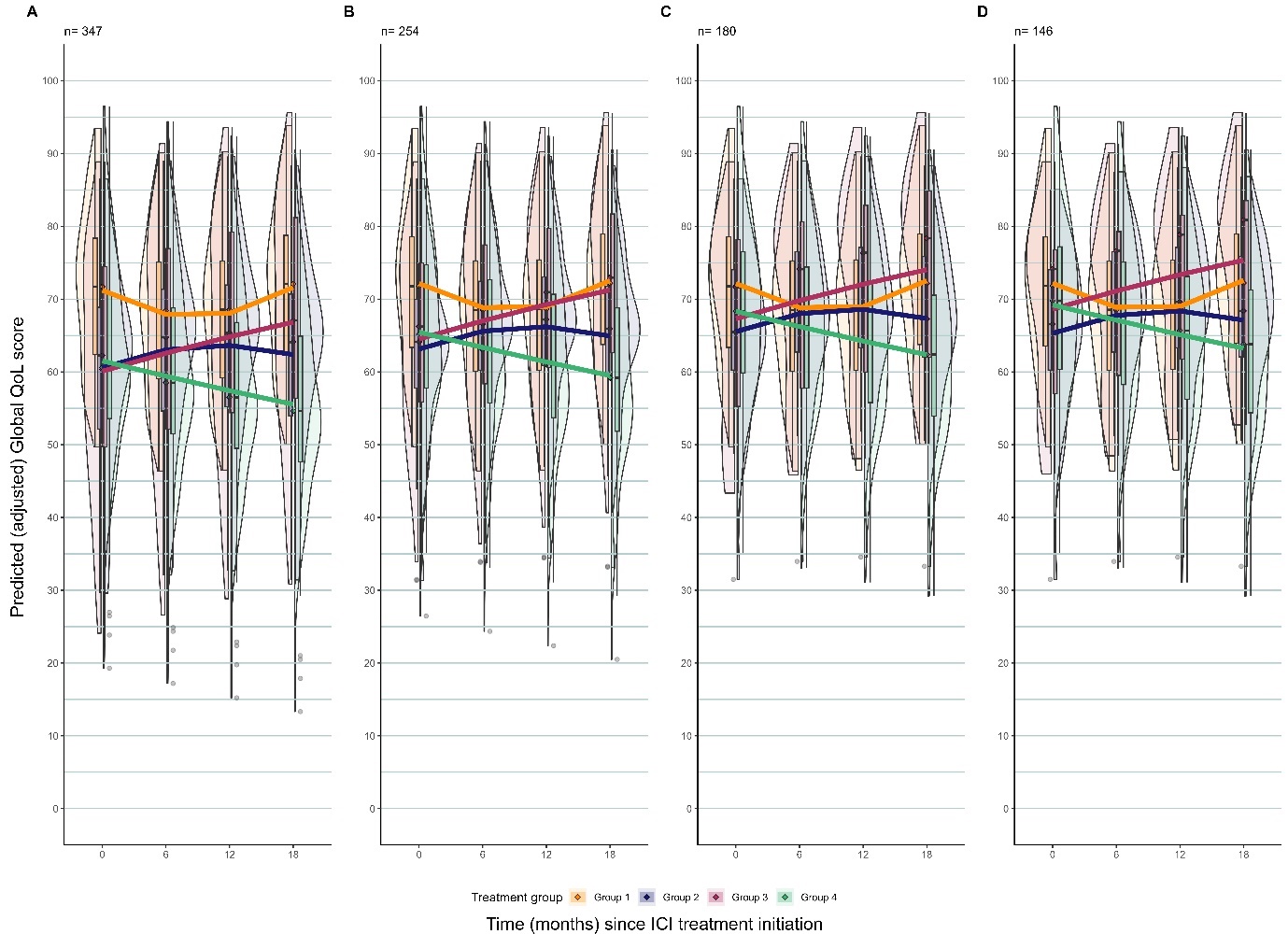

**Figure A.9**: Predicted trajectory of Global QoL scores from a Joint model by treatment group with ‘observation time’ as a time-dependant continuous covariate and adjusted for pre-specified covariates and competing risk of death using a Cox-PH model (complete case analysis). An interaction term between treatment group and ‘observation time’ and ‘observation time’ squared is included in the model. Violin plots of the subject-level predicted (adjusted) gQoL score are superimposed with a curve showing the population-level predicted (adjusted) gQoL score for A. the full cohort, B. those who survived at least 6 months, C. those who survived at least 12 months, D. those who survived at least 18 months. Panel A. also shows all other population level-predicted (adjusted) gQoL scores from panels B-D on a single graph depicting the relative difference in the trajectories among these cohorts.

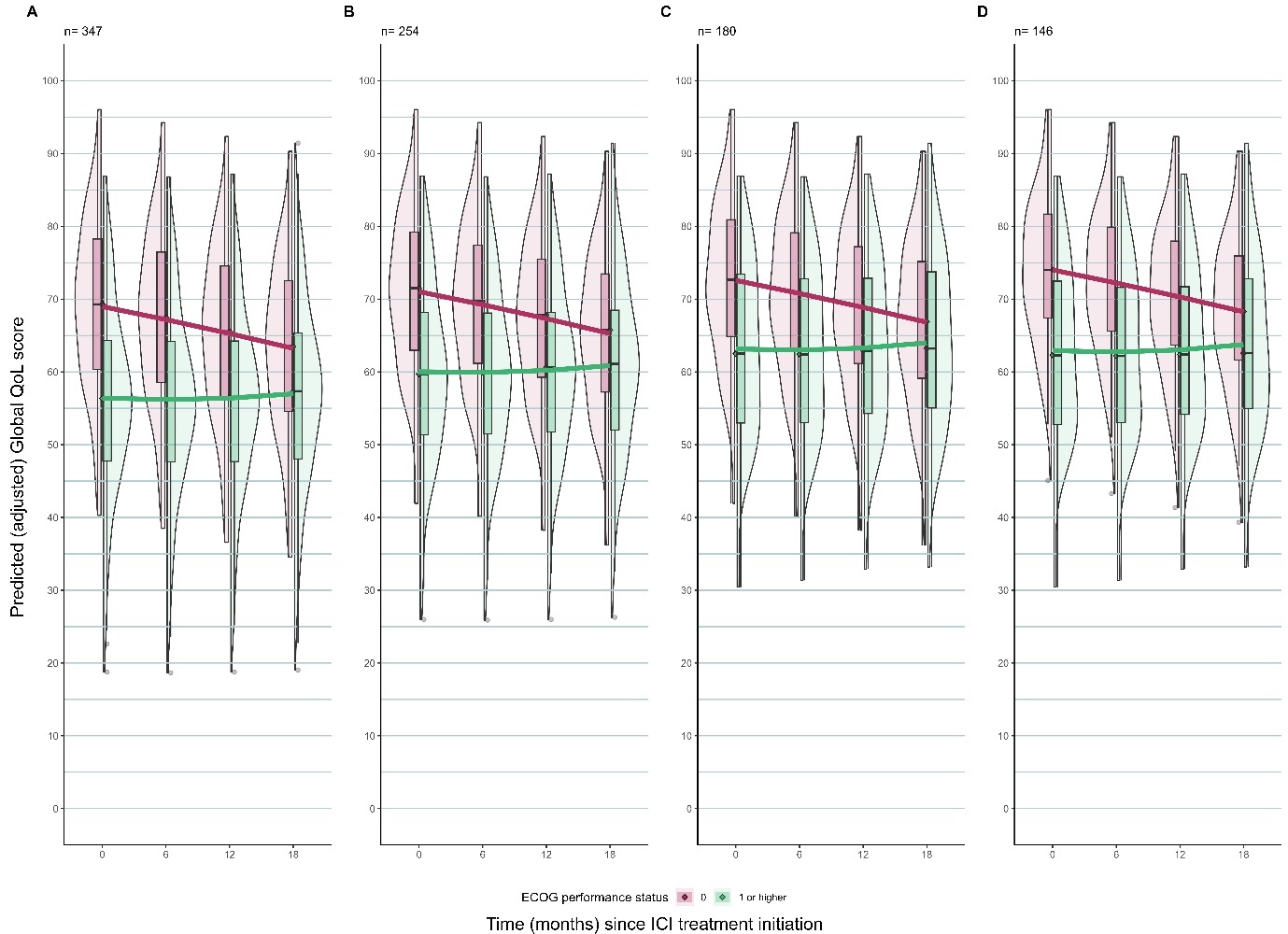

**Figure A.10**: Predicted trajectory of Global QoL scores from a Joint model by ECOG PS with ‘observation time’ as a time-dependant continuous covariate and adjusted for pre-specified covariates and competing risk of death using a Cox-PH model (complete case analysis). An interaction term between ECOG PS and ‘observation time’ and ‘observation time’ squared is included in the model. Violin plots of the subject-level predicted (adjusted) gQoL score are superimposed with a curve showing the population-level predicted (adjusted) gQoL score for A. the full cohort, B. those who survived at least 6 months, C. those who survived at least 12 months, D. those who survived at least 18 months. Panel A. also shows all other population level-predicted (adjusted) gQoL scores from panels B-D on a single graph depicting the relative difference in the trajectories among these cohorts.

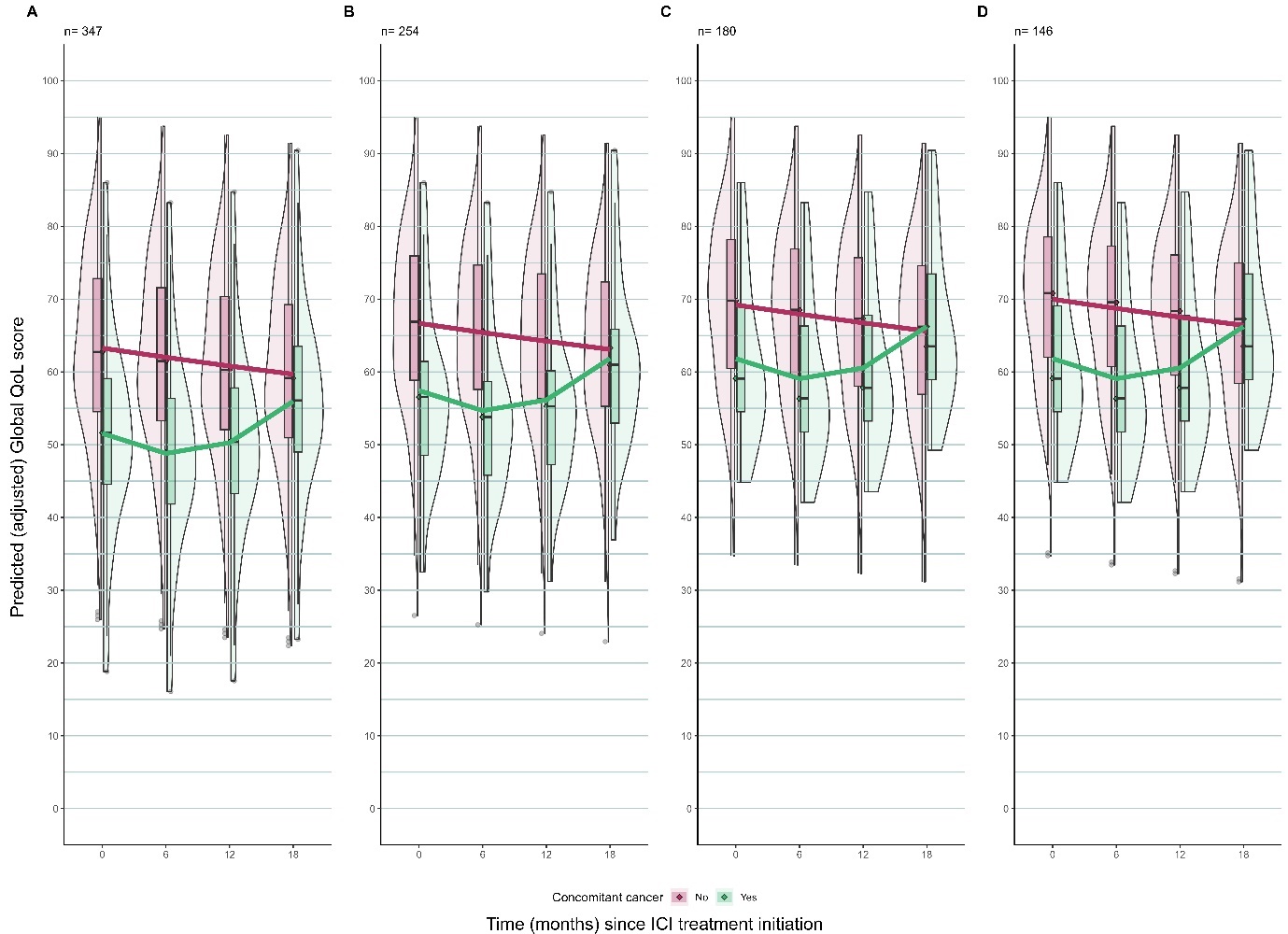

**Figure A.11**: Predicted trajectory of Global QoL scores from a Joint model by presence of concomitant cancer with ‘observation time’ as a time-dependant continuous covariate and adjusted for pre-specified covariates and competing risk of death using a Cox-PH model (complete case analysis). An interaction term between concomitant cancer and ‘observation time’ and ‘observation time’ squared is included in the model. Violin plots of the subject-level predicted (adjusted) gQoL score are superimposed with a curve showing the population-level predicted (adjusted) gQoL score for A. those who survived at least 6 months, C. those who survived at least 12 months, D. those who survived at least 18 months. Panel A. also shows all other population level-predicted (adjusted) gQoL scores from panels B-D on a single graph depicting the relative difference in the trajectories among these cohorts.

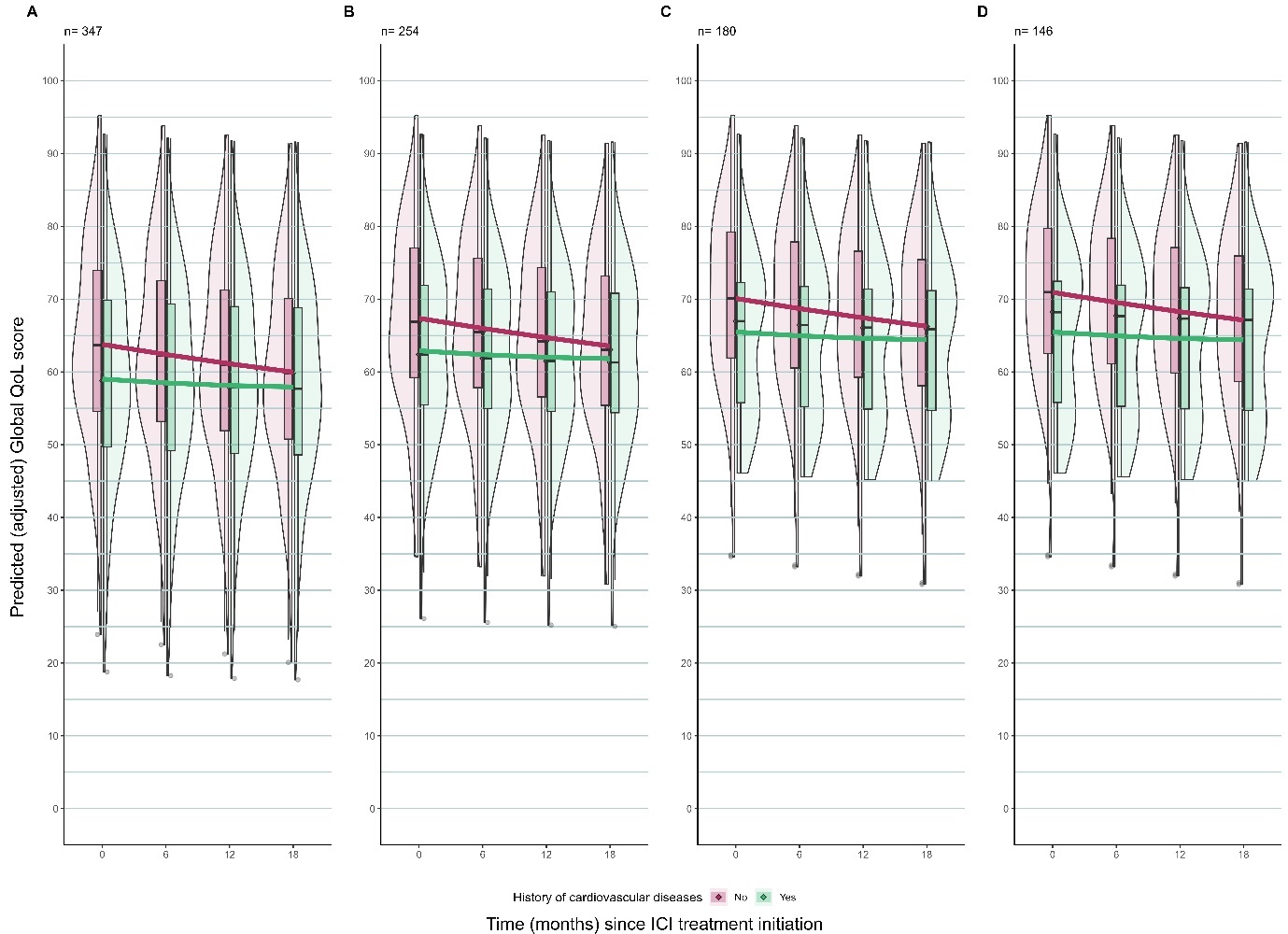

**Figure A.12**: Predicted trajectory of Global QoL scores from a Joint model by history of CVD with ‘observation time’ as a time-dependant continuous covariate and adjusted for pre-specified covariates and competing risk of death using a Cox-PH model (complete case analysis). An interaction term between history of CVD and ‘observation time’ and ‘observation time’ squared is included in the model. Violin plots of the subject-level predicted (adjusted) gQoL score are superimposed with a curve showing the population-level predicted (adjusted) gQoL score for A. the full cohort, B. those who survived at least 6 months, C. those who survived at least 12 months, D. those who survived at least 18 months. Panel A. also shows all other population level-predicted (adjusted) gQoL scores from panels B-D on a single graph depicting the relative difference in the trajectories among these cohorts.

**Appendix 5**

**Extended results from joint models with multiple imputation**

**Model description**

The outcome here was the gQoL which was measured repeatedly over time, first at baseline and then every 6 months to up to 18 months. The exposure was a categorical variable of treatment groups, where group 1 consists of patients who had first line chemo-radiotherapy followed by durvalumab within 6 weeks to 3 months with a curative intent, group 2 consists of patients who had first line immuno-monotherapy with a palliative intent and no other prior anti-cancer treatments, group 3 consists of patients who had first line immune-chemotherapy with a palliative intent and group 4 consists of patients who had second or further line immunotherapy (either alone or in combination with chemotherapy) with a palliative intent.. The covariables adjusted in the analysis were age, sex, weight, education, PS, comorbidities, concomitant cancer, tumour stage and number of months since cancer diagnosis. Observation time for QoL measurements and time-to-death since ICI treatment initiation were coded as continuous variables on the same scale (years). A quadratic term for time of QoL measurement (observation time) was added to reflect the shape of the (unadjusted) relationship between QoL score and observation time (Figure 1). To facilitate model convergence and interpretation, variables age and weight were standardised (centred at their mean). The gQoL score was scaled by dividing by 100 so it lies between 0 and 1.

Convergence of Monte Carlo Markov Chains (MCMC) were assessed by trace plots, which displayed horizontal bands across chains, and the Gelman-Rubin criterion for convergence was satisfied. The Monte Carlo error was not more than 5% of the standard deviation of posterior mean.

**Table A.3:** Posterior summary from Bayesian linear mixed effect sub-model of the joint model with multiple imputation

| **Variable** |  | **95 % CI of PM** | |
| --- | --- | --- | --- |
|  | **PM** | **2.50%** | **97.50%** |
| Intercept | 0.62 | 0.50 | 0.73 |
| Treatment group 1 ( vs group 4) | 0.10 | -0.02 | 0.23 |
| Treatment group 2 ( vs group 4) | 0.06 | 0.00 | 0.11 |
| Treatment group 3 ( vs group 4) | 0.04 | -0.03 | 0.11 |
| Weight | 0.02 | 0.00 | 0.04 |
| ECOG PS = 1 (vs PS = 0) | -0.09 | -0.13 | -0.05 |
| ECOG PS = 2 (vs PS = 0) | -0.09 | -0.18 | 0.00 |
| ECOG PS = 3 (vs PS = 0) | -0.09 | -0.32 | 0.14 |
| Diabetes (vs no) | -0.01 | -0.06 | 0.05 |
| Have hypertension (vs no) | 0.03 | -0.01 | 0.07 |
| Have COPD (vs no) | 0.02 | -0.03 | 0.06 |
| Have rheumatological conditions (vs no) | -0.01 | -0.08 | 0.06 |
| Have history of CVD (vs no) | -0.05 | -0.10 | -0.01 |
| Have concomitant cancer (vs no) | -0.09 | -0.17 | -0.02 |
| Age | 0.00 | -0.03 | 0.02 |
| Female sex | 0.02 | -0.03 | 0.07 |
| Stage 4 (vs Stage = 3) | 0.01 | -0.11 | 0.12 |
| Medium-level Education (vs Low-level) | -0.04 | -0.09 | 0.02 |
| High-level Education (vs Low-level) | 0.00 | -0.05 | 0.06 |
| Number of months since lung cancer diagnosis | 0.01 | -0.02 | 0.04 |
| Observation time for QoL | 0.05 | -0.01 | 0.12 |
| (Observation time for QoL)^2^ | -0.04 | -0.08 | 0.00 |

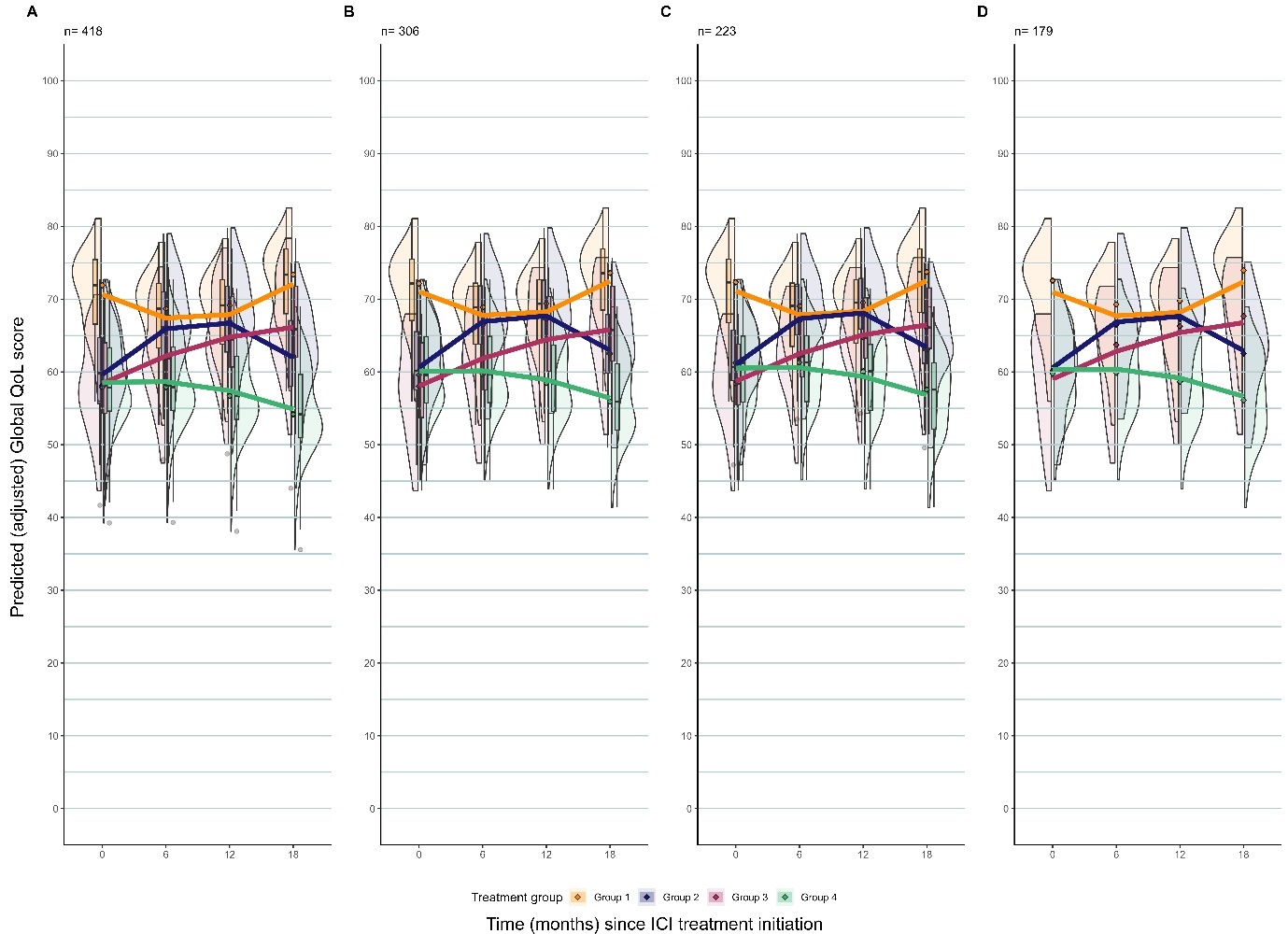

**Figure A.13**: Predicted trajectory of Global QoL scores from a Joint model with multiple imputation by treatment group with ‘observation time’ as a time-dependant continuous covariate and adjusted for pre-specified covariates and competing risk of death using a Cox-PH model. An interaction term between treatment group and ‘observation time’ and ‘observation time’ squared is included in the model. Violin plots of the subject-level predicted Global QoL score with superimposed curves showing the population-level predicted Global QoL score by treatment group for A) the full cohort B) those who survived at least 6 months C) those who survived at least 12 months and D) those who survived at least 18 months.

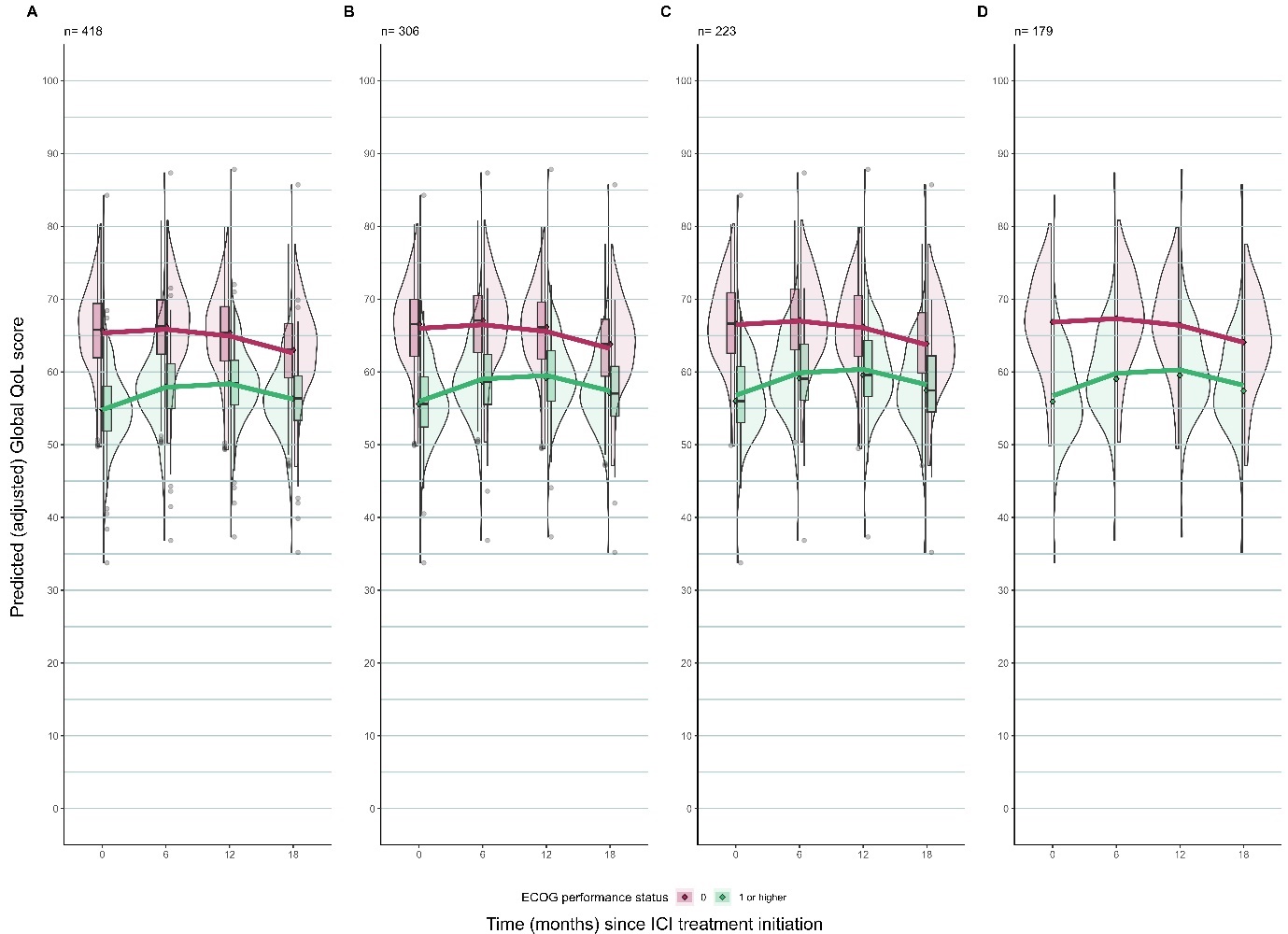

**Figure A.14**: Predicted trajectory of Global QoL scores from a Joint model with multiple imputation by ECOG PS with ‘observation time’ as a time-dependant continuous covariate and adjusted for pre-specified covariates and competing risk of death using a Cox-PH model. An interaction term between PS and ‘observation time’ and ‘observation time’ squared is included in the model. Violin plots of the subject-level predicted Global QoL score with superimposed curves showing the population-level predicted Global QoL score by treatment group for A) the full cohort B) those who survived at least 6 months C) those who survived at least 12 months and D) those who survived at least 18 months.

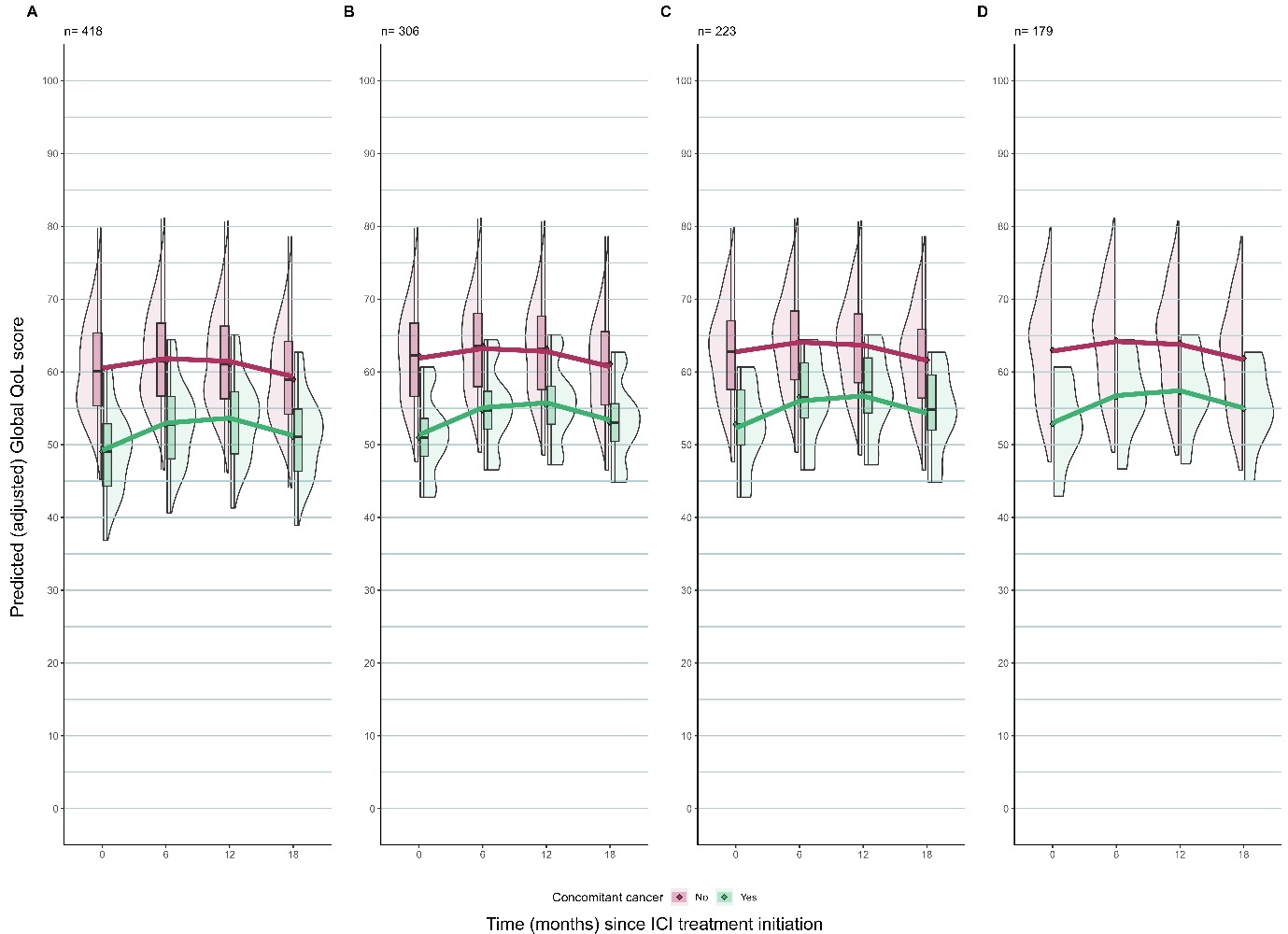

**Figure A.15**: Predicted trajectory of Global QoL scores from a Joint model with multiple imputation by presence of concomitant cancer with ‘observation time’ as a time-dependant continuous covariate and adjusted for pre-specified covariates and competing risk of death using a Cox-PH model. An interaction term between concomitant cancer and ‘observation time’ and ‘observation time’ squared is included in the model. Violin plots of the subject-level predicted Global QoL score with superimposed curves showing the population-level predicted Global QoL score by treatment group for A) the full cohort B) those who survived at least 6 months C) those who survived at least 12 months and D) those who survived at least 18 months.

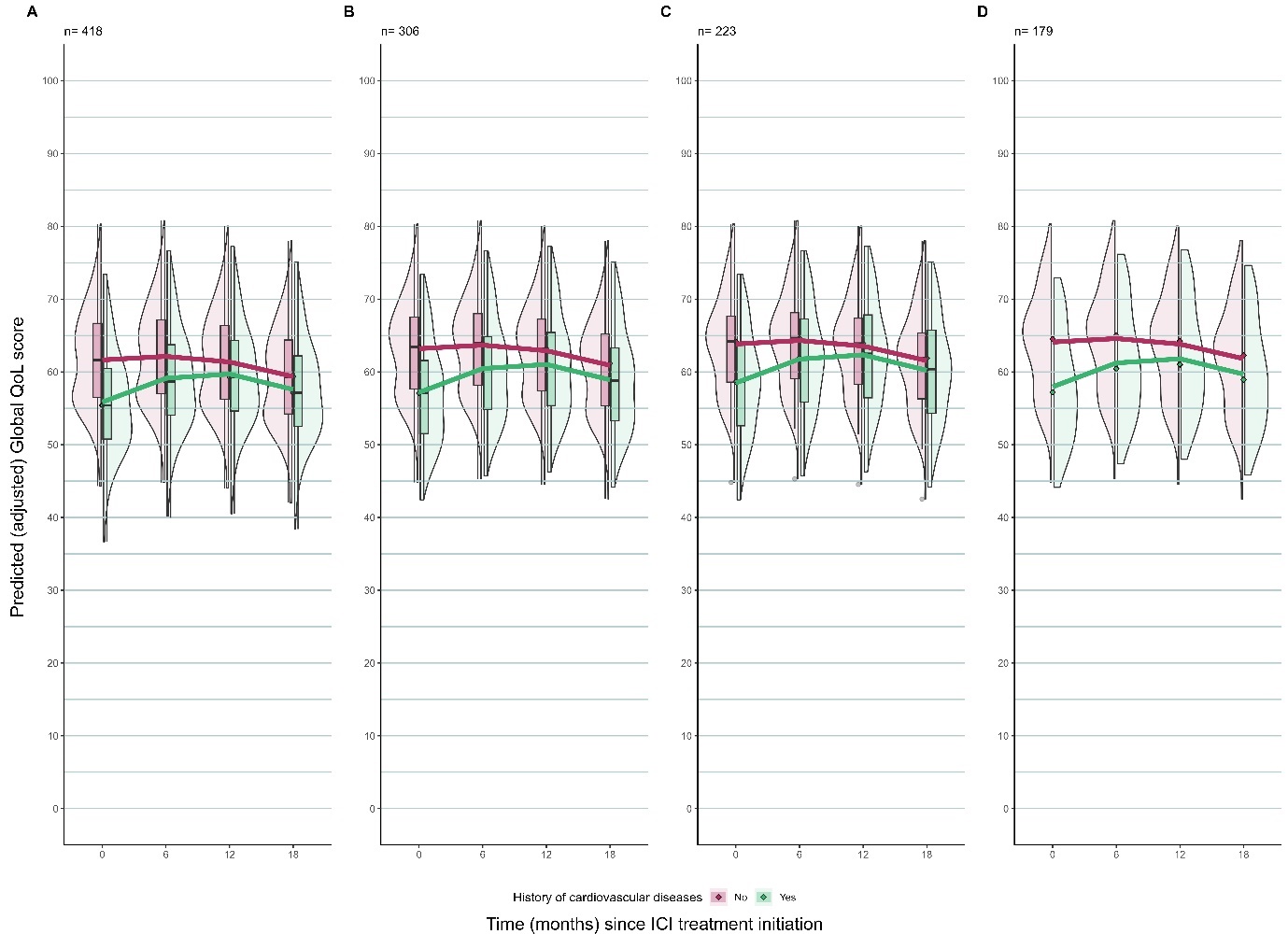

**Figure A.16**: Predicted trajectory of Global QoL scores from a Joint model with multiple imputation by history of CVD cancer with ‘observation time’ as a time-dependant continuous covariate and adjusted for pre-specified covariates and competing risk of death using a Cox-PH model. An interaction term between CVD and ‘observation time’ and ‘observation time’ squared is included in the model. Violin plots of the subject-level predicted Global QoL score with superimposed curves showing the population-level predicted Global QoL score by treatment group for A) the full cohort B) those who survived at least 6 months C) those who survived at least 12 months and D) those who survived at least 18 months.

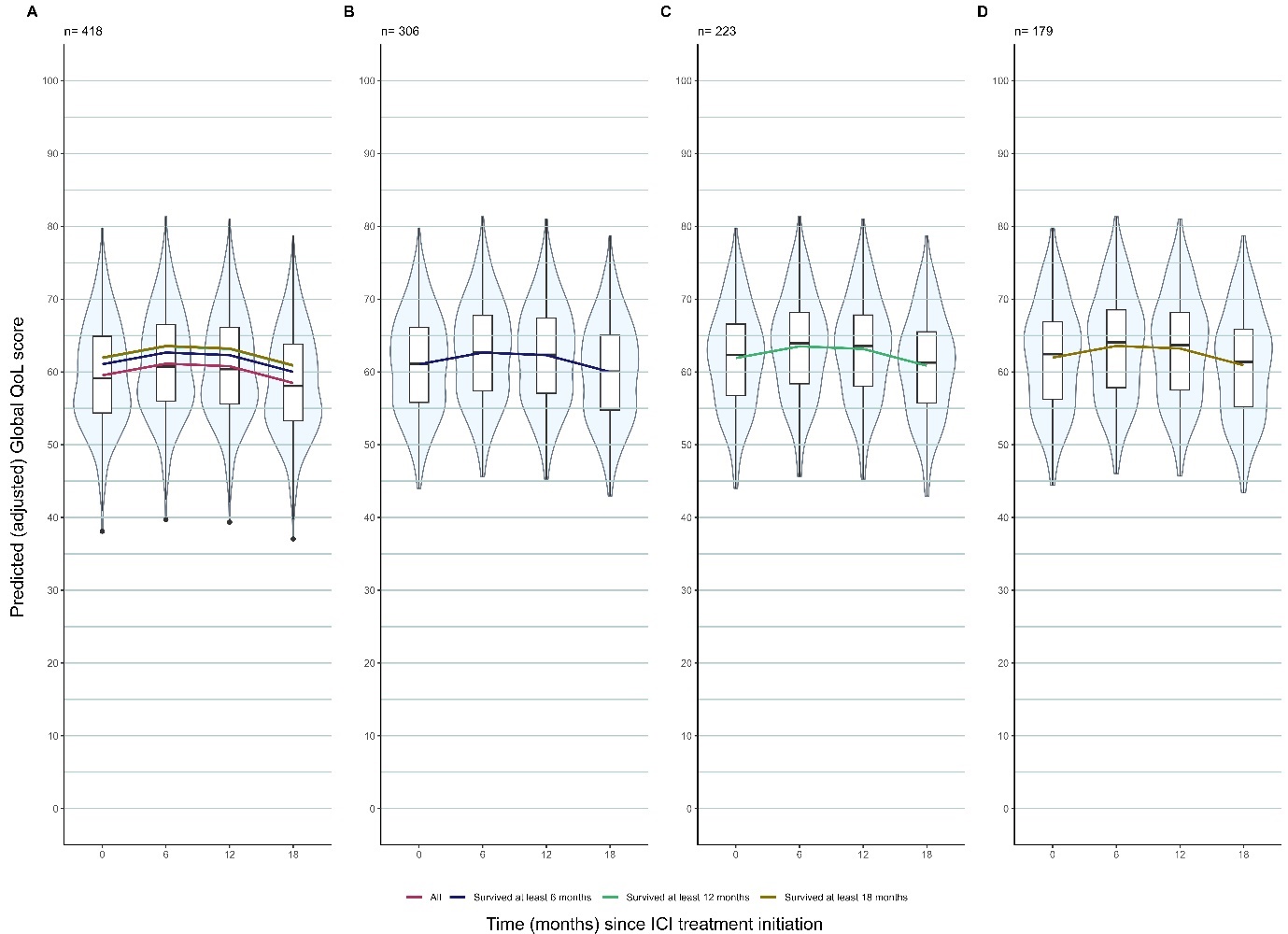

**Figure A.17:** Predicted trajectory of gQoL scores of patients using joint model with multiple imputation accounting for the competing risk of death (sensitivity analysis). Violin plots summarise the subject-level predicted (adjusted) gQoL score and are superimposed with a curve showing the population-level predicted (adjusted) gQoL score for A. the full cohort, B. those who survived at least 6 months, C. those who survived at least 1 months, D. those who survived at least 18 months. Panel A. also shows all the population level-predicted (adjusted) gQoL scores from panels B-D on a single graph depicting the relative difference in the trajectories among these cohorts by survival time.

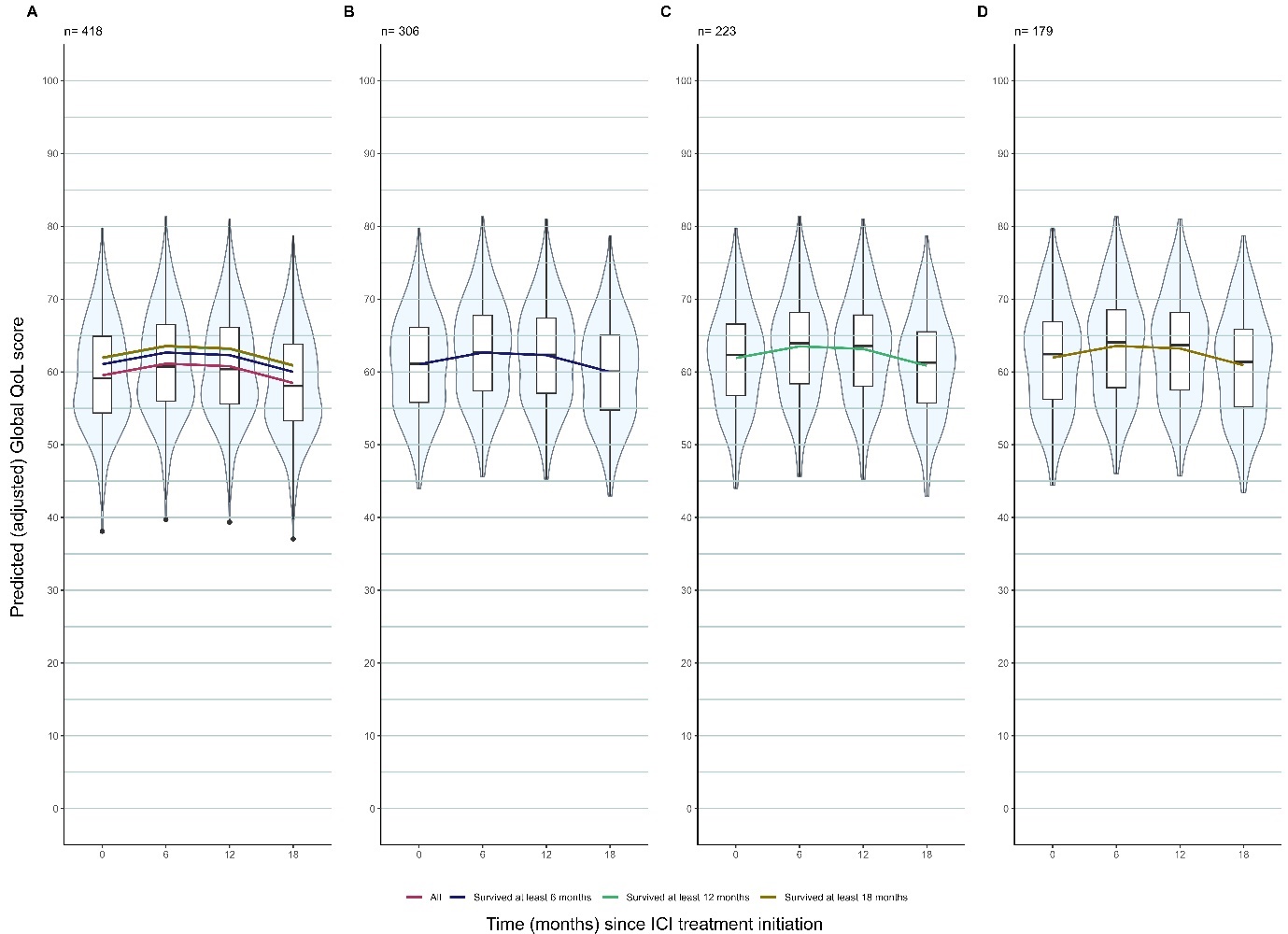

**Figure A.18:** Predicted trajectory of gQoL scores of patients using joint model with multiple imputation accounting for the competing risk of death (sensitivity analysis). Covariables with missing values are weight and PS. Missing values on education are imputed with education = low. Violin plots of the subject-level predicted (adjusted) gQoL score are superimposed with a curve showing the population-level predicted (adjusted) gQoL score for A. the full cohort, B. those who survived at least 6 months, C. those who survived at least 1 months, D. those who survived at least 18 months. Panel A. also shows all the population level-predicted (adjusted) gQoL scores from panels B-D on a single graph depicting the relative difference in the trajectories among these cohorts.

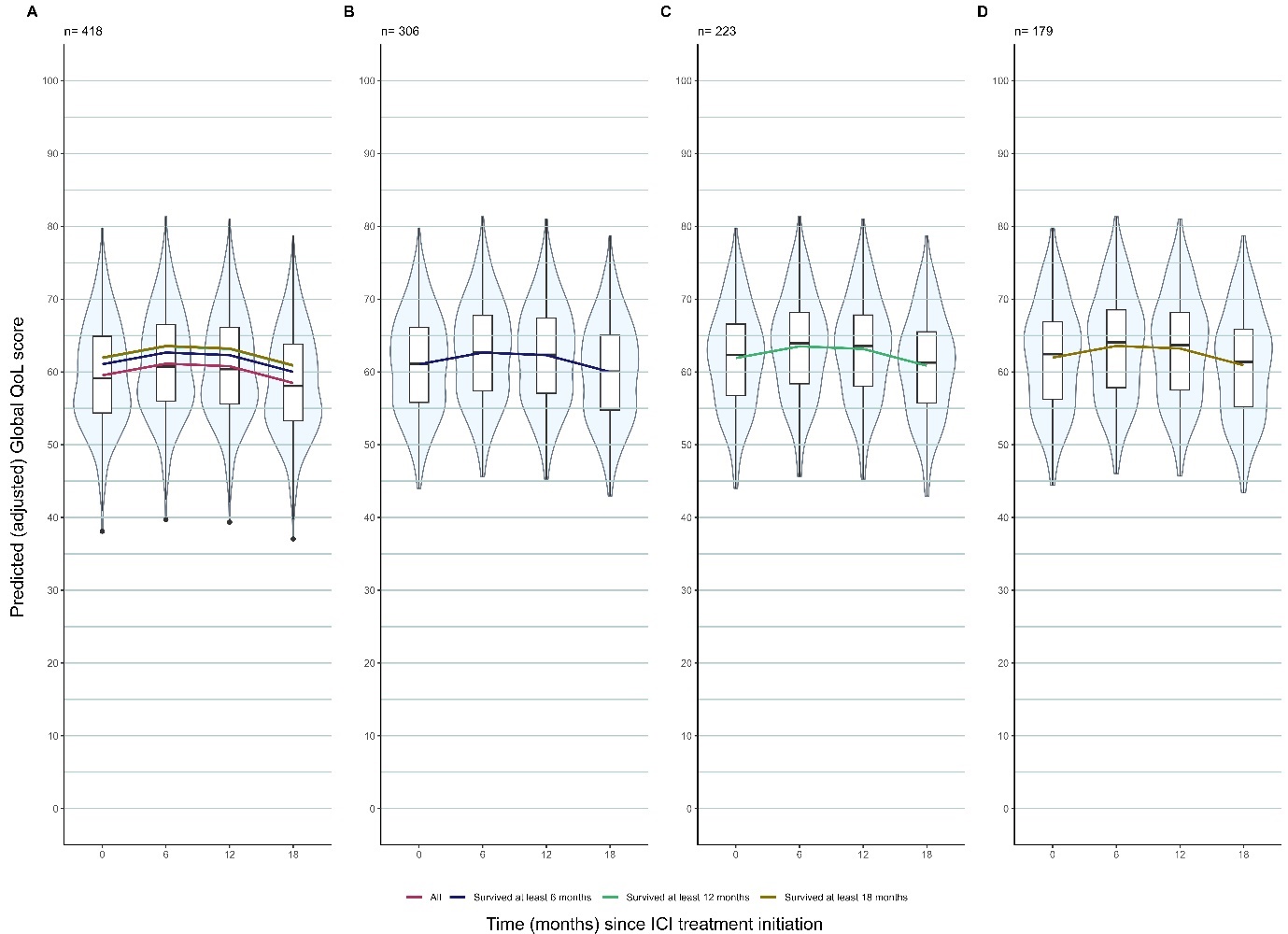
 **Figure A.19:** Predicted trajectory of gQoL scores of patients using joint model with multiple imputation accounting for the competing risk of death (sensitivity analysis). Covariables with missing values are weight and PS. Missing values on education are imputed with education = high. Violin plots of the subject-level predicted (adjusted) gQoL score are superimposed with a curve showing the population-level predicted (adjusted) gQoL score for A. the full cohort, B. those who survived at least 6 months, C. those who survived at least 1 months, D. those who survived at least 18 months. Panel A. also shows all the population level-predicted (adjusted) gQoL scores from panels B-D on a single graph depicting the relative difference in the trajectories among these cohorts.
